# Transcranial Photobiomodulation on Language and Cognitive Performance in Down Syndrome: A Pilot Randomized Sham-Controlled Trial

**DOI:** 10.64898/2026.07.03.26357051

**Authors:** Fabio Luchese, Puneet Velidi, Layane Robert Bou Jaoude, Lauren Sidelinger, Maia Gersten, Borja Ignacio Ferreras, Carlos Lohmann, Aura Maria Hurtado Puerto, Julie A. Clancy, Kayla McEachern, Katelyn Sylvester, Suk-tak Chan, Margaret Pulsifer, Margaret Naeser, Anita Saltmarche, Elizabeth Corcoran, Angela John Thurman, Leonard Abbeduto, Brian G. Skotko, Paolo Cassano

**Affiliations:** Division of Neuromodulation and Neuropsychiatry, Massachusetts General Hospital, Boston, MA, USA; Department of Psychiatry, Harvard Medical School, Boston, MA, USA; Department of Mathematics and Statistics, University of Victoria, Victoria, BC, Canada; Department of Psychiatry, Massachusetts General Hospital, Boston, MA, USA; VA Boston Healthcare System, Boston, MA, USA; Department of Neurology, Boston University School of Medicine, Boston, MA, USA; Saltmarche Health and Associates, Toronto, ON, Canada; Down’s Syndrome Research Foundation, London, UK; MIND Institute and Department of Psychiatry and Behavioral Sciences, University of California Davis Health, Sacramento, CA, USA; Down Syndrome Program, Division of Medical Genetics and Metabolism, Department of Pediatrics, Massachusetts General Hospital, Boston, MA, USA; Department of Pediatrics, Harvard Medical School, Boston, MA, USA

**Keywords:** Down syndrome, transcranial photobiomodulation, near-infrared light, EEG gamma power, language, grammar omissions, neuromodulation, pilot randomized trial

## Abstract

**Background:** Down syndrome (DS) is associated with persistent language and cognitive impairments and with abnormalities in cortical oscillatory activity, including in the gamma range. Transcranial photobiomodulation (tPBM) is a noninvasive neuromodulatory intervention with potential benefits for cortical physiology, language, and cognition.

**Methods:** We conducted a pilot randomized, double-blind, sham-controlled trial of repeated 40-Hz near-infrared tPBM in adolescents and young adults with DS. Fourteen participants were randomized 1:1 to active tPBM or sham and received 18 sessions over 6 weeks, followed by short-term and long-term follow-up. Outcomes included resting-state EEG gamma power, connected-speech measures, language and cognitive indices, and selected computerized tasks.

**Results:** Active tPBM did not significantly increase global resting-state EEG gamma power relative to sham at either follow-up. Pre-registered analyses did not show broad treatment benefit across outcomes, although they did identify a significant short-term advantage for active tPBM on grammatical morpheme accuracy in connected speech; in contrast, picture naming favored sham at long-term follow-up. Exploratory mechanistic analyses did not show a robust biological treatment signal. Both active and sham procedures were well tolerated, with no serious adverse events.

**Conclusions:** In this underpowered pilot sample, 6 weeks of 40-Hz near-infrared tPBM–delivered unilaterally, at low power, on target areas–did not demonstrate a meaningful effect size in DS. A dose-finding study for tPBM in DS, also accounting for age of participants, is recommended.

## Introduction

Down syndrome (DS) is the most common genetic cause of intellectual disability and is associated with both lifelong neurodevelopmental vulnerability and accelerated brain aging (Hart et al., 2017; Lott and Head, 2019). Improved medical care has extended survival into adulthood, increasing the clinical importance of persistent language and cognitive difficulties as well as age-related neurodegeneration (Wu and Morris, 2013; Hart et al., 2017). Effective interventions targeting language and cognition in DS remain limited (Hart et al., 2017). Triplication of chromosome 21, including increased amyloid precursor protein (APP) dosage, contributes to the neurodevelopmental phenotype of DS and to the markedly elevated risk of Alzheimer’s disease (AD), with AD neuropathology present in nearly all adults with DS by midlife (Wiseman et al., 2015; Lott and Head, 2019).

Language impairment is one of the most clinically meaningful features of DS. Expressive language is typically more affected than receptive language or nonverbal ability, with prominent weaknesses in morphosyntax, intelligibility, and connected speech (Martin et al., 2009). These challenges affect daily communication and make language a relevant therapeutic target. Measures such as narrative production, verbal fluency, naming, and articulation provide clinically meaningful probes of distributed language systems (Martin et al., 2009).

A neurophysiological rationale for intervention comes from evidence linking DS to altered cortical excitability and abnormal oscillatory activity, including in the gamma range (Buzsáki and Wang, 2012; Ruiz-Mejias et al., 2016; Iaccarino et al., 2016; Martorell et al., 2019). Gamma oscillations support local circuit computation and cognition, and DS models show impaired gamma activity related to altered interneuron function (Buzsáki and Wang, 2012; Ruiz-Mejias et al., 2016). In AD models, 40-Hz stimulation can reduce amyloid burden and improve disease-relevant phenotypes, while human EEG studies in DS report abnormalities across alpha-, beta-, and gamma-range activity (Iaccarino et al., 2016; Martorell et al., 2019; Hamburg et al., 2021; Velikova et al., 2011).

Transcranial photobiomodulation (tPBM) is a noninvasive neuromodulatory approach using red or near-infrared light to influence cortical physiology (Hamblin, 2016). NIR light can reach superficial cortex, and PBM has been linked to mitochondrial, cerebrovascular, oxygenation, and electrophysiological effects (Tedford et al., 2015; Hamblin, 2016; Saucedo et al., 2021; Vargas et al., 2017; Zomorrodi et al., 2019). Although clinical effects vary by wavelength, dose, pulsing, montage, and population, meta-analytic evidence supports cognitive effects in healthy adults, and preliminary aphasia studies suggest possible language-related benefits (Salehpour et al., 2019; Naeser et al., 2020).

Despite this rationale, DS-specific evidence for tPBM remains limited. The present pilot randomized, double-blind, sham-controlled study was designed to test whether repeated 40-Hz NIR tPBM could modulate resting-state EEG gamma power and improve language performance in adolescents and young adults with DS, as well as have positive effects on attention and memory. In addition, we assessed whether treatment-related changes in gamma activity were associated with changes in language and cognitive outcomes and explored possible effects on resting-state functional connectivity. We hypothesized that active tPBM, relative to sham, would produce greater short-term and longer-term improvements across neurophysiological, language, and cognitive domains.

## Materials and Methods

### Study Design

This pilot randomized, double-blind, sham-controlled, parallel-group trial evaluated the effects of repeated 40-Hz near-infrared transcranial photobiomodulation in adolescents and young adults with Down syndrome. The study was conducted at Massachusetts General Hospital/Mass General Brigham and was registered at ClinicalTrials.gov (NCT04668001). The protocol was approved by the local institutional review board, and written informed consent was obtained from each participant’s legal guardian, with participant assent obtained when appropriate.

Potential participants underwent an initial pre-screening followed by an in-person or remote screening visit. Baseline assessments included neuropsychological testing, a standardized narrative language sample, resting-state EEG, and optional resting-state functional MRI (rs-fMRI). After completion of baseline assessments, participants were randomized in a 1:1 ratio to active tPBM or sham. Participants, caregivers, treatment staff, and outcome assessors were blinded to treatment assignment. Blinding integrity was assessed during treatment using the Perceived Blinding Questionnaire, administered at sessions 9 and 18. Participants received 18 treatment sessions over 6 weeks, followed by short-term and long-term follow-up assessments. The short-term follow-up occurred within approximately 1 week after the final treatment session, and the long-term follow-up occurred approximately 4 weeks later. An open-label extension was available per protocol, however it is not the focus of the present report (Figure 1).

**Figure 1.**
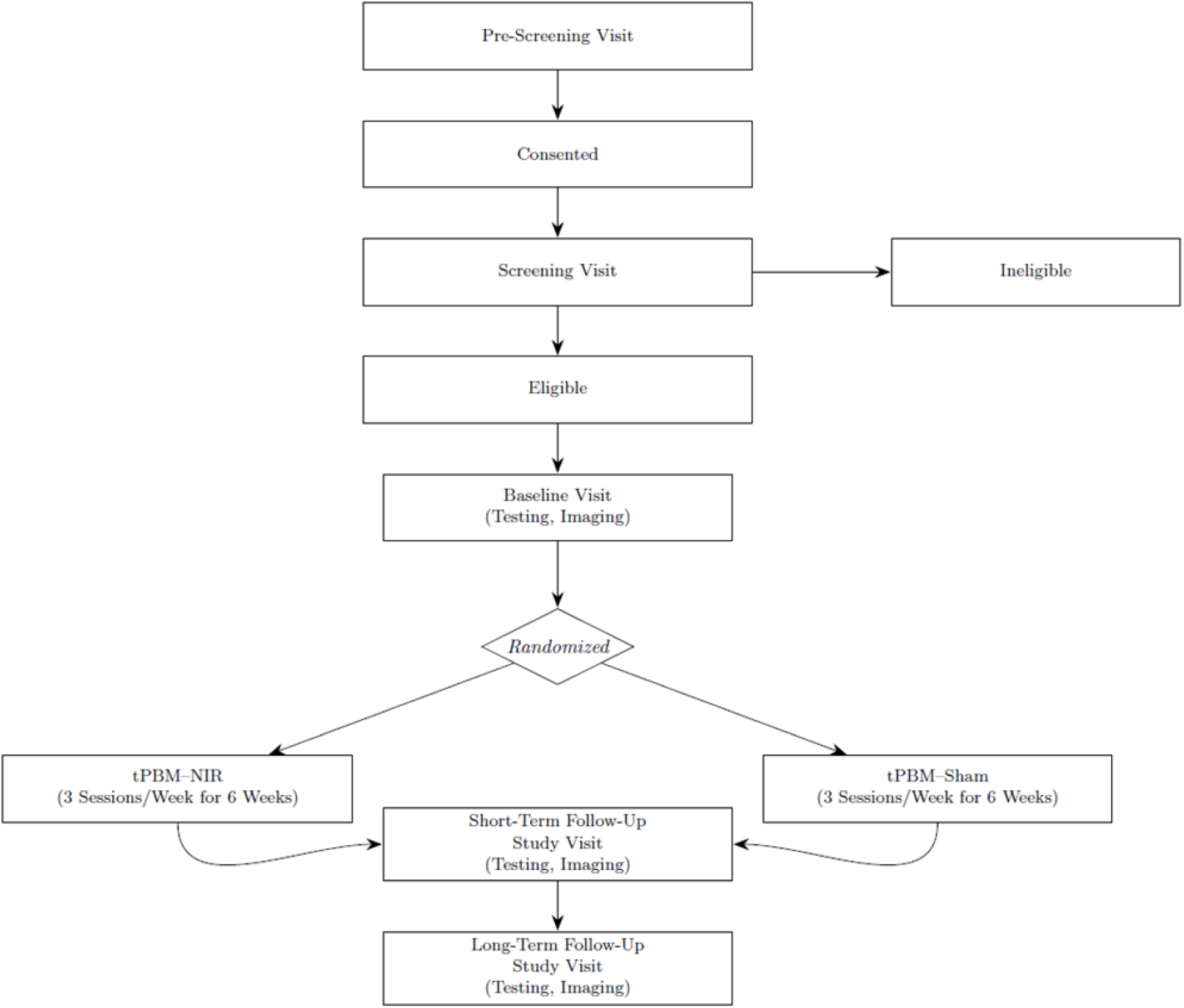
Study schema and assessment schedule. Pilot randomized, double-blind, sham-controlled, parallel-group trial of 40-Hz near-infrared transcranial photobiomodulation (tPBM) in adolescents and young adults with Down syndrome. After screening and baseline assessments, eligible participants were randomized 1:1 to active tPBM or sham and received 18 treatment sessions over 6 weeks (3 sessions/week). Study assessments were performed at baseline, short-term follow-up (within approximately 1 week after the final treatment session), and long-term follow-up (approximately 4 weeks after the final treatment session). Language and cognitive assessments and resting-state EEG were collected longitudinally; resting-state fMRI was optional. Open-label treatment was available per protocol but is not included in the present report.

### Participants

Eligibility criteria included age 16–35 years, documented diagnosis of DS, and sufficient spoken English skills to complete the language assessments (i.e., production of at least occasional multiword utterances). Participants were encouraged to attend with a caregiver, to support adherence and to enable caregiver-reported outcomes. Exclusion criteria encompassed seizure disorder, dementia, inability to complete study procedures, primary reliance on non-speech communication or spontaneous speech of fewer than 2-word utterances, recent or anticipated changes in medications or therapies at the investigator’s discretion, untreated obstructive sleep apnea, participation in another interfering clinical trial, current pregnancy, active cancer treatment, migraine with aura within the previous 6 months, and conditions increasing risk from NIR exposure. MRI participation was optional; participants with MRI contraindications or unwillingness to undergo MRI remained eligible for all other study procedures.

### tPBM intervention

Active and sham interventions were delivered using the MedX 1100 system (MedX Health Corp, Mississauga, ON) (Figure 2). In the active condition, stimulation was delivered at a wavelength of 870 nm in pulsed mode at 40 Hz with a 50% duty cycle (Table 1). Sessions lasted 30 minutes and were administered 3 times weekly for 6 weeks, for a total of 18 sessions. Five stimulation sites were targeted simultaneously: mesial prefrontal cortex, precuneus, left inferior frontal/perisylvian cortex (Broca’s area), left temporal cortex (Wernicke’s area region), and, when feasible, the left angular gyrus (Table 2). If head size precluded placement of the fifth diode, that site was omitted while maintaining the same fluence per active site.

**Figure 2.**
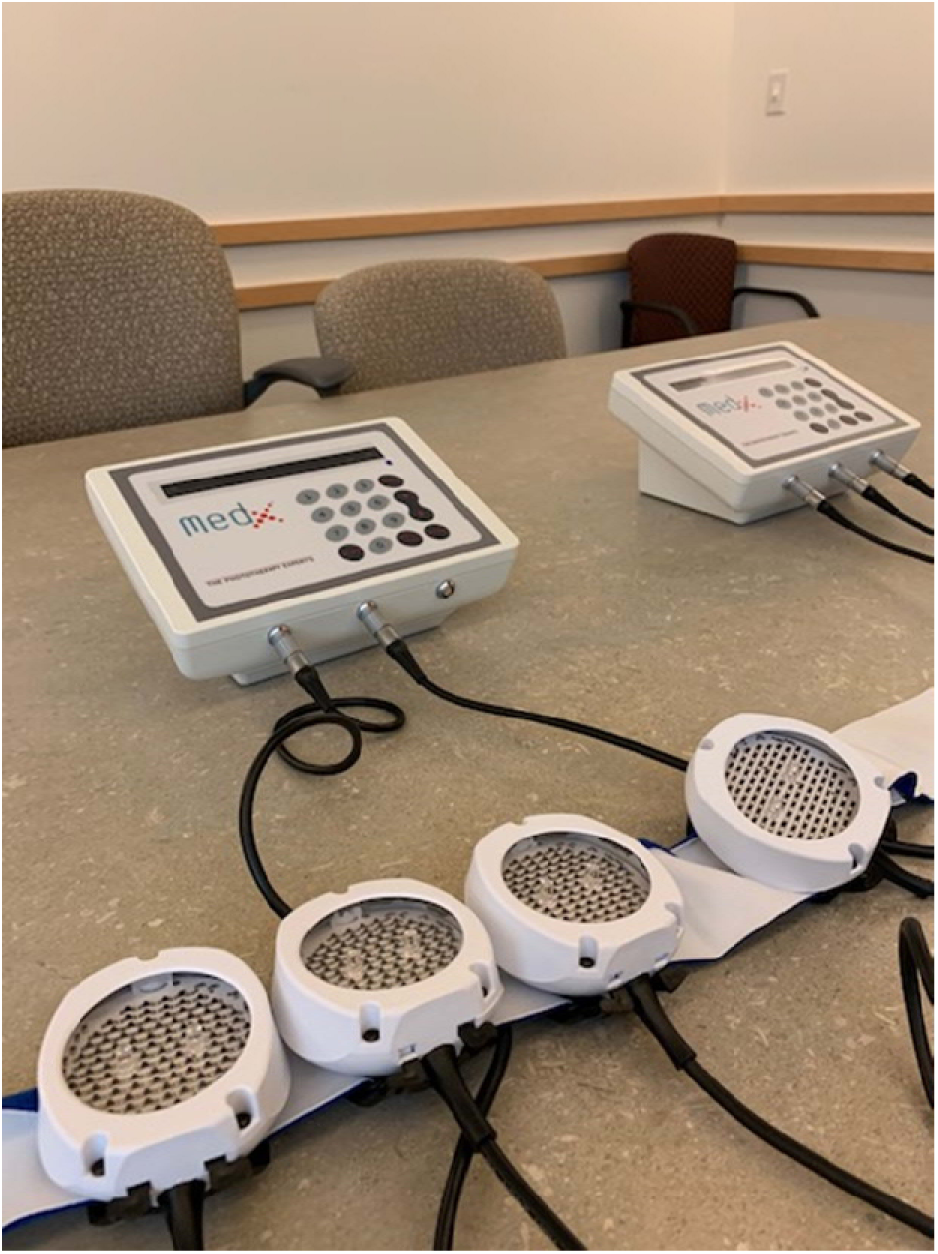
**tPBM–NIR device (MedX 1100) with LED cluster heads.**

**Table 1.** tPBM Parameters.

| Measure (unit) | Value |
| --- | --- |
| Wavelength (nm) | 870, near-infrared (NIR) |
| Pulse rate (Hz) | 40 |
| Duty cycle | 50% |
| Exposure time | 30 min per session, delivered simultaneously across 5 sites |
| Window/spot size (cm<sup>2</sup>) | 22.48 cm<sup>2</sup> per site (diameter 5.345 cm) |
| Total area exposed (cm<sup>2</sup>) | ~112 cm<sup>2</sup> (22.48 × 5) |
| Irradiance / power density (mW/cm<sup>2</sup>) | 44.48 |
| Average Irradiance | 22.24 |
| Fluence delivered per LED cluster head (J/cm<sup>2</sup>) | 40 |
| Power per LED cluster head (mW) | 1,000 |
| Device | MedX 1100 LED cluster heads, used simultaneously |

**Note 1.** Fluence was calculated as irradiance × time × duty cycle. With an irradiance of 44.48 mW/cm<sup>2</sup> (0.04448 W/cm<sup>2</sup>) and a 50% duty cycle, delivery of 40 J/cm<sup>2</sup> requires approximately 1800 s (30 min) per site: $40 / (0.04448 \times 0.5) \approx 1799$ s. Without pulsing (100% duty cycle), delivery of 1 J/cm<sup>2</sup> would require approximately 22.5 s.
**Note 2.** To improve tolerability, participants could be titrated from 20 J/cm<sup>2</sup> to 40 J/cm<sup>2</sup> during the first few treatments. If 40 J/cm<sup>2</sup> was not tolerated, treatment could be continued at 20 or 30 J/cm<sup>2</sup>.

**Table 2.** tPBM Device Placements.

| Placement | Target brain area |
| --- | --- |
| Center of the forehead, at the junction with the frontal hairline | Mesial prefrontal cortex |
| Junction of the midsagittal and lambdoid suture lines | Precuneus cortex |
| Left temple | Broca's area |
| Above the left ear (T3) | Wernicke's area / left temporal cortex |
| Posterior and superior to T3 | Left angular gyrus area (BA39) |

**Note.** The fifth diode (left angular gyrus site) was omitted when head size did not allow placement of all five diodes.

Each LED cluster head delivered an irradiance of 44.48 mW/cm² over 22.48 cm², corresponding to a fluence of 40 J/cm² per site during a 30-minute session. To improve tolerability, dosing could be titrated from 20 J/cm² to 40 J/cm² during the first few sessions; if 40 J/cm² was not tolerated, treatment could continue at 20 or 30 J/cm². The sham device was identical in appearance and procedures and generated mild warmth but no measurable NIR output at the scalp. Protective eyewear was used at all sessions (although not strictly necessary, given the light sources were LEDs).

### Clinical Outcome Measures

Clinical outcome measures were selected to address the prespecified language and attention/memory aims. All assessments were administered by trained, blinded raters.

Language outcomes included the Expressive Language Sampling (ELS) Wordless Picture Book narrative task, in which connected-speech samples were audio-recorded, transcribed, and coded to derive transcript-based measures of lexical diversity, intelligibility, mean length of utterance in morphemes, and grammatical morpheme omissions (Thurman et al., 2021); the RBANS language subtests, including Picture Naming and Semantic Fluency (Randolph et al., 1998); and the Goldman-Fristoe Test of Articulation–Third Edition (GFTA-3) (Goldman and Fristoe, 2015).

Attention and memory outcomes included RBANS Attention subtests (Digit Span and Coding), RBANS Memory subtests (List Learning, List Recall, and List Recognition) (Randolph et al., 1998), and selected CANTAB tasks: Reaction Time, Paired Associates Learning, and Motor Screening (Fray and Robbins, 1996).

Score types and directionality of improvement for each outcome are summarized in Supplementary Table 1.

General cognitive ability was characterized at baseline using the Kaufman Brief Intelligence Test– Second Edition (KBIT-2) IQ Composite standard score (Kaufman and Kaufman, 2004).

### EEG acquisition and processing

Resting-state EEG was acquired primarily with the Neuroelectrics StarStim 32-channel system and, when that system was unavailable, with the Advanced Brain Monitoring StatX24 20-channel system. The StarStim system used a linked right mastoid reference and electrodes positioned according to the international 10–20 system; the StatX24 system used a standard linked-mastoid montage. EEG was recorded during both eyes-open and eyes-closed resting-state conditions, with approximately 5 minutes acquired per condition; longer or repeated acquisitions were allowed when needed to obtain usable data.

Raw EEG data were processed in MNE-Python. Gamma-band activity was quantified from the 35–45 Hz band-pass-filtered signal as mean squared amplitude, averaged across all channels and timepoints to yield a single gamma-power value per recording. Gamma power was log10-transformed prior to statistical analysis to improve interpretability and stabilize variance. Recordings were summarized at baseline, post-treatment, and follow-up.

### MRI acquisition and exploratory functional connectivity analysis

MRI data were acquired on a 3T scanner with multichannel receiver coils using protocols developed at the Martinos Center for Biomedical Imaging. When feasible, participants underwent high-resolution 3D T1-weighted MPRAGE, resting-state functional MRI, and a diffusion tensor imaging (DTI) sequence. For the exploratory resting-state functional connectivity analyses, we use the Glasser360 Atlas with Yeo7 ROI network definitions to define prespecified regions of interest, including language-related cortical regions (Broca’s area, Wernicke’s area, and mid-temporal cortex, and the supramarginal and angular gyrus areas) as well as canonical network nodes from the Default Mode Network (medial prefrontal cortex, precuneus, posterior cingulate cortex, angular gyrus, and lateral parietal cortex) For exploratory rs-fMRI analyses, data were preprocessed using the longitudinal fMRIPrep pipeline (v24) and postprocessed in XCP-D with a 36-parameter denoising strategy. Scans were excluded if more than 50% of BOLD volumes had framewise displacement >0.5 mm. One participant was excluded because incomplete T1 acquisition precluded adequate preprocessing. Given the limited and inconsistent availability of fMRI data across sessions, rsFC findings were treated as exploratory.

### Statistical analysis

Analyses were conducted in R version 4.3.2. Data were screened for missing or incomplete assessments, temporal inconsistencies, and out-of-range values, with discrepancies resolved against source records.

For the pre-registered primary analyses, two change scores were computed for each outcome: Δ1 = short-term follow-up (STF) − baseline and Δ2 = long-term follow-up (LTF) − baseline. Interpretation of Δ was scale-dependent: positive Δ values indicated improvement for higher-is-better outcomes, whereas negative Δ values indicated improvement for lower-is-better outcomes, including CANTAB latency/error measures, GFTA-3 raw articulation error scores, and grammatical morpheme omissions from the ELS Wordless Picture Book task.

Between-group comparisons were conducted separately at each timepoint using Welch’s t-test when assumptions were approximately met and Mann–Whitney U tests otherwise. Effect sizes were reported as Hedges’ g for parametric tests and rank-biserial r for non-parametric tests, with 95% confidence intervals. No multiplicity adjustment was applied across the full primary outcome battery because of the pilot, estimation-focused nature of the study. Primary change-score analyses were complete-case at each timepoint, without imputation.

Supportive baseline-adjusted linear mixed-effects models were fit for clinical outcomes using the form Outcome ∼ Group × Time + baseline + (1|Participant), with sham as the reference group and time coded as STF and LTF. Estimated marginal means and prespecified between-group contrasts were obtained using emmeans. Inference used Satterthwaite degrees of freedom and cluster-robust CR2 standard errors, with Holm adjustment across the two follow-up contrasts within each outcome. Models used all available observations without imputation.

For EEG, change in log10-transformed gamma power from baseline to post-treatment and follow-up was analyzed using linear mixed-effects models with fixed effects for treatment, session, and their interaction, and a random intercept for participant. Supportive Bayesian analyses estimated the posterior probability that active-treatment change exceeded sham. Exploratory EEG–clinical outcome associations were restricted to the active tPBM group and used Pearson correlations and simple linear regression models relating gamma change to changes in language and cognitive outcomes; these analyses were not corrected for multiple comparisons.

Exploratory rs-fMRI treatment effects on functional connectivity were assessed using linear mixed-effects models with random intercept for participant and fixed effect for treatment group, with supportive Bayesian analyses using uninformative priors. Given the small imaging subsample and multiple regional comparisons, rs-fMRI results were interpreted cautiously.

Baseline demographic and clinical characteristics were summarized descriptively by treatment group. Descriptive between-group comparisons used Welch’s t-test for continuous variables and Fisher’s exact tests for categorical variables because of the small sample size and sparse cell counts.

## Results

### Participant flow and baseline characteristics

A total of 19 individuals were formally screened/consented. Of these, 5 did not proceed to randomization: 3 were unable to complete the required study procedures/tasks, primarily because of insufficient ability to comply with language and/or cognitive task instructions, and 2 withdrew or declined to proceed. Thus, 14 participants were enrolled and randomized in a 1:1 ratio to active tPBM or sham (7 per group), and all randomized participants completed the study procedures. Clinical and behavioral outcome analyses were based on complete-case data at each timepoint, with supportive mixed-effects models using all available observations without imputation. For resting-state EEG, analyzable data were available for 13 participants (active tPBM n = 7; sham n = 6). Resting-state fMRI was optional per protocol; because of limited and inconsistent scan availability across sessions, and because one participant was excluded due to incomplete T1 acquisition precluding preprocessing, the exploratory rs-fMRI analyses were based on a small imaging subsample (active tPBM n = 3; sham n = 4) (Figure 3).

**Figure 3.**
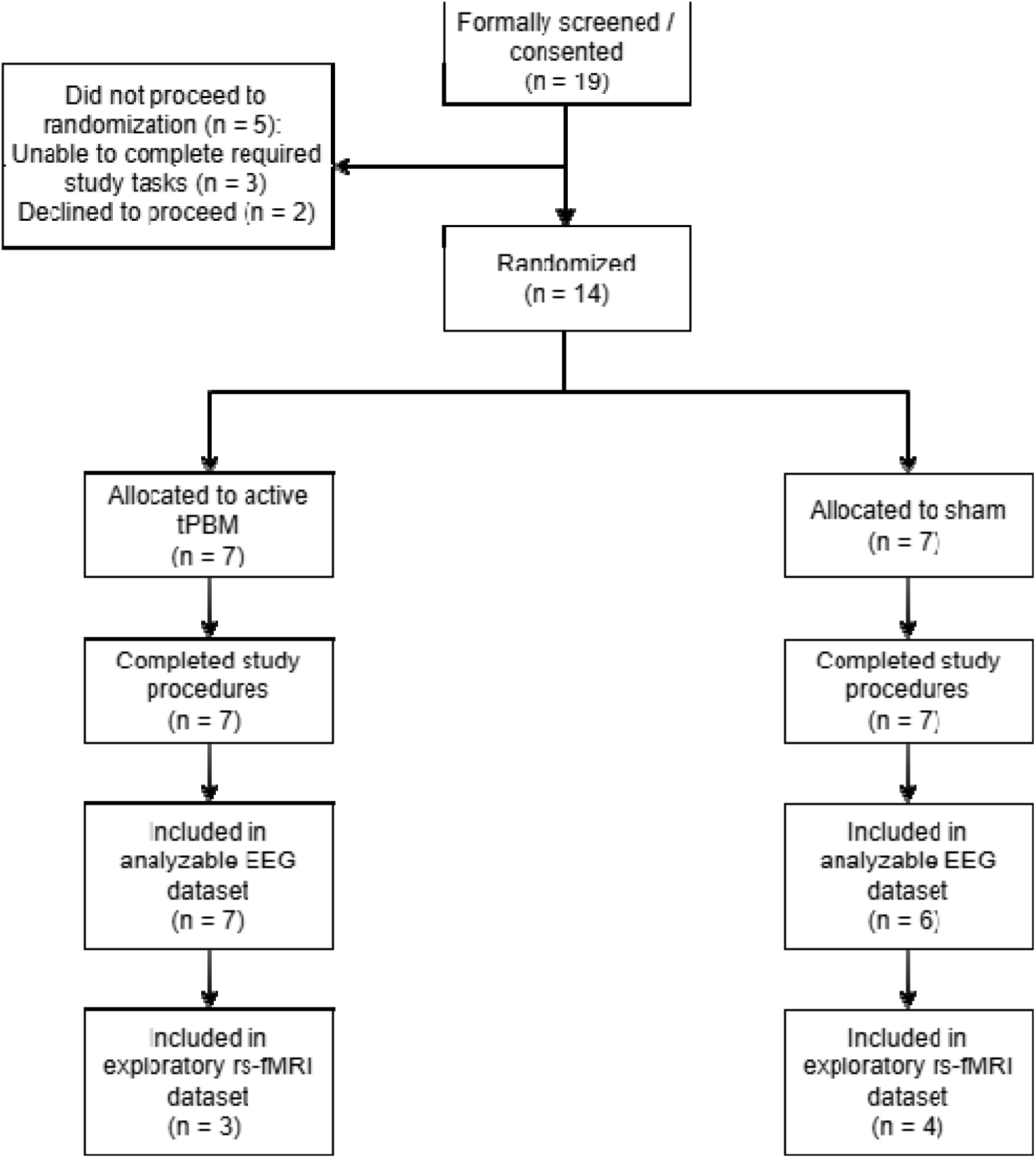
CONSORT flow diagram and analytic samples. Flow of participants from screening through randomization, study completion, and inclusion in the EEG and exploratory rs-fMRI analyses. All randomized participants completed study procedures. Denominators differ across analytic modalities because rs-fMRI was optional per protocol and some datasets were not available or analyzable after preprocessing/quality-control procedures.

Baseline demographic and clinical characteristics are summarized in Table 3. The mean age of the overall sample was 24.7 ± 5.5 years (range, 17.4–34.9), with similar mean age across the two groups. Baseline medical comorbidities and concomitant treatments were heterogeneous, as expected in a young cohort with DS. Documented psychiatric conditions—based on reported medical history—were present in 6/14 participants (42.9%), equally distributed across groups, and included depression, anxiety, ADHD, OCD, or behavioral modification. Of note, in case of medical or psychiatric comorbidity, only subjects with stable conditions were allowed into the study. Psychotropic medication use was observed in 5/14 participants (35.7%), and psychotherapy/behavioral support in 2/14 (14.3%). Some numerical baseline imbalances were observed, including sex distribution, but descriptive between-group comparisons did not identify statistically significant baseline differences.

**Table 3.**
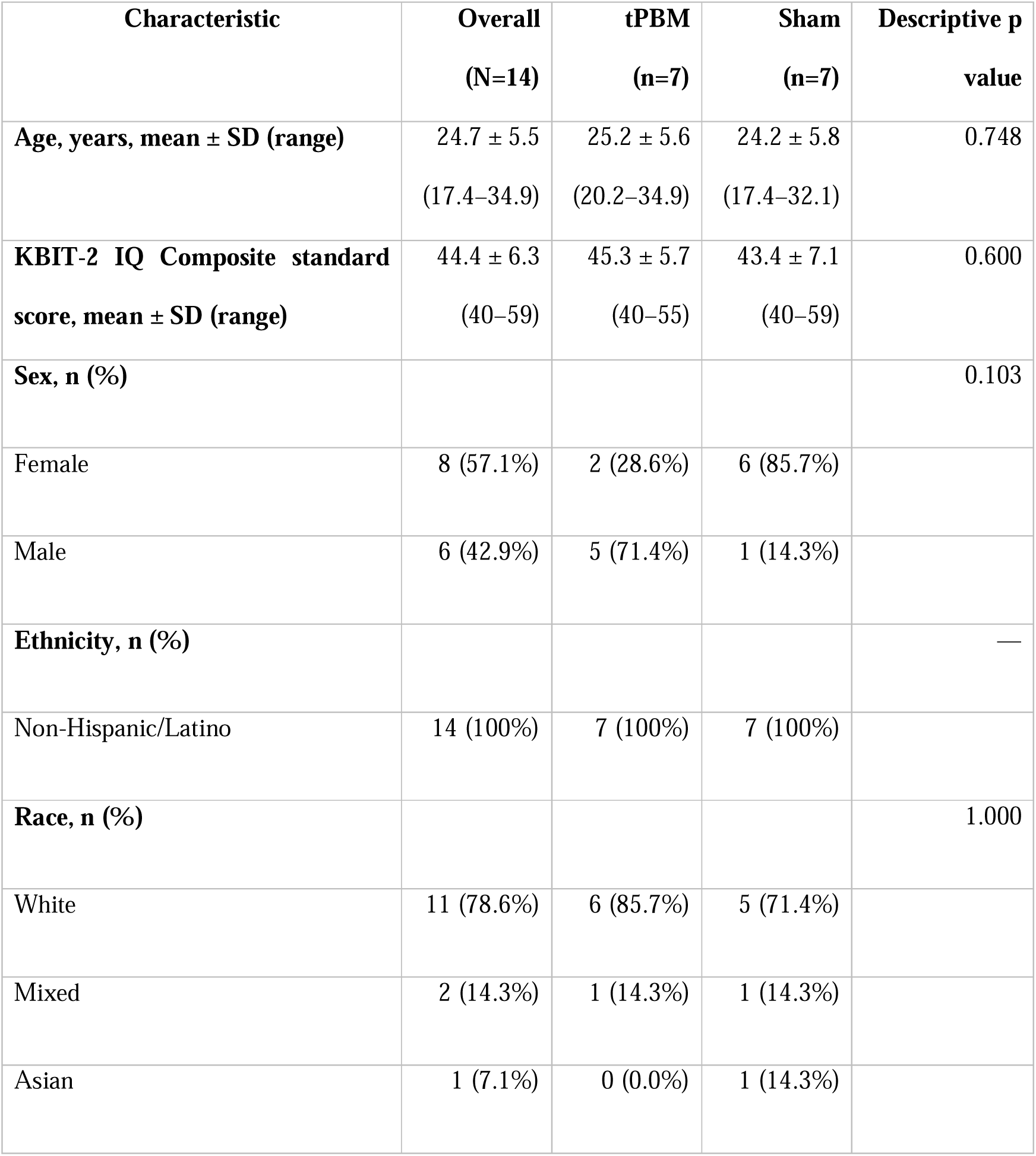
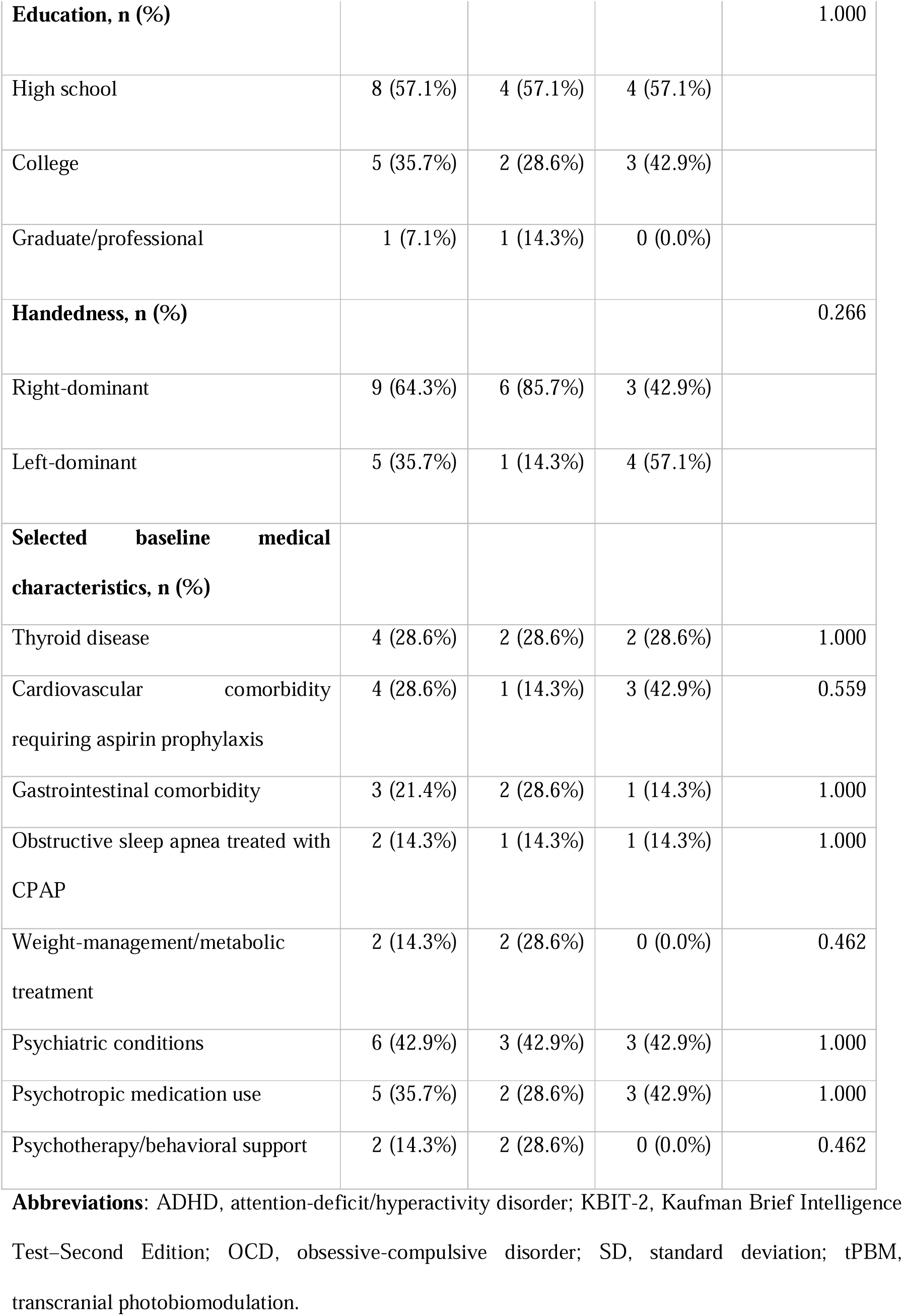
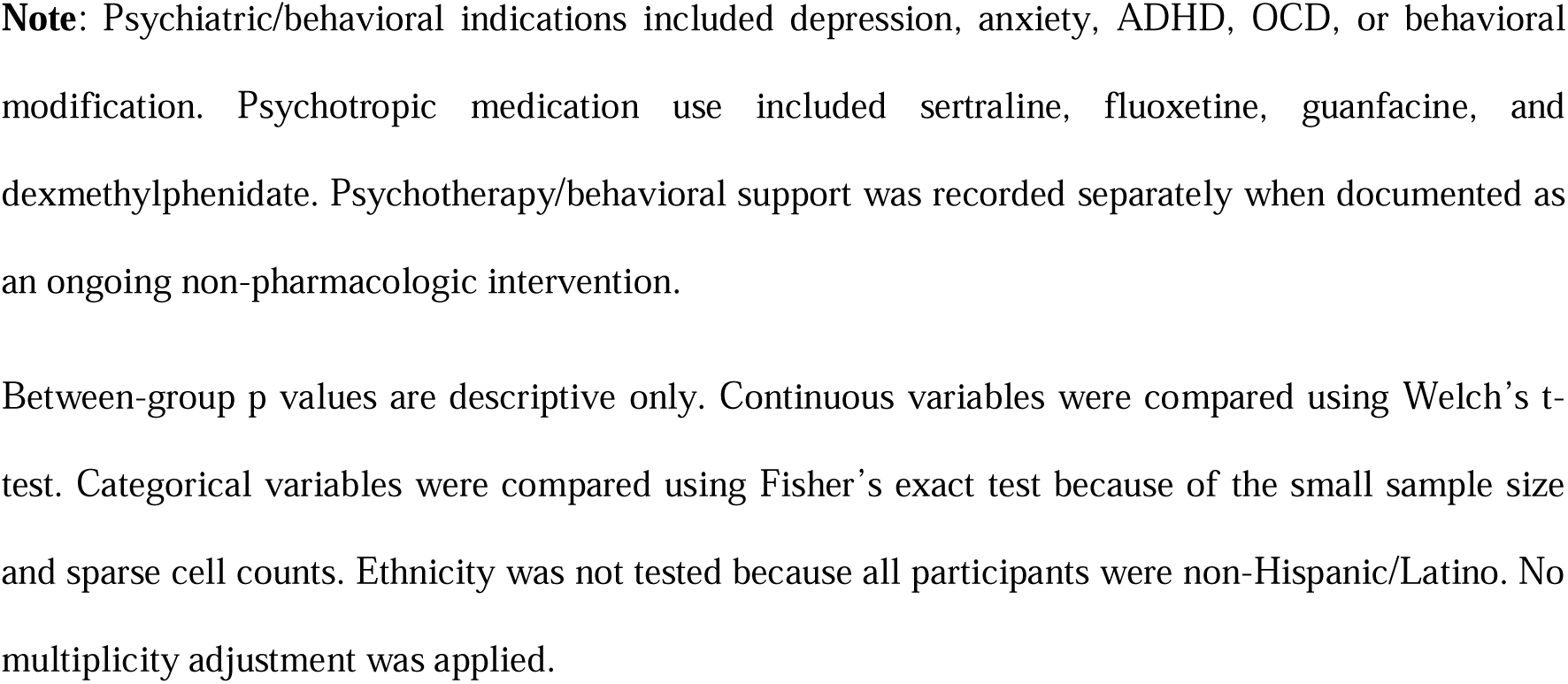
Baseline characteristics of study participants.

| **Characteristic** | **Overall**<br>**(N=14)** | **tPBM**<br>**(n=7)** | **Sham**<br>**(n=7)** | **Descriptive p**<br>**value** |
| --- | --- | --- | --- | --- |
| **Age, years, mean ± SD (range)** | 24.7 ± 5.5<br>(17.4–34.9) | 25.2 ± 5.6<br>(20.2–34.9) | 24.2 ± 5.8<br>(17.4–32.1) | 0.748 |
| **KBIT-2 IQ Composite standard score, mean ± SD (range)** | 44.4 ± 6.3<br>(40–59) | 45.3 ± 5.7<br>(40–55) | 43.4 ± 7.1<br>(40–59) | 0.600 |
| **Sex, n (%)** |  |  |  | 0.103 |
| Female | 8 (57.1%) | 2 (28.6%) | 6 (85.7%) |  |
| Male | 6 (42.9%) | 5 (71.4%) | 1 (14.3%) |  |
| **Ethnicity, n (%)** |  |  |  | — |
| Non-Hispanic/Latino | 14 (100%) | 7 (100%) | 7 (100%) |  |
| **Race, n (%)** |  |  |  | 1.000 |
| White | 11 (78.6%) | 6 (85.7%) | 5 (71.4%) |  |
| Mixed | 2 (14.3%) | 1 (14.3%) | 1 (14.3%) |  |
| Asian | 1 (7.1%) | 0 (0.0%) | 1 (14.3%) |  |
| **Education, n (%)** |  |  |  | 1.000 |
| High school | 8 (57.1%) | 4 (57.1%) | 4 (57.1%) |  |
| College | 5 (35.7%) | 2 (28.6%) | 3 (42.9%) |  |
| Graduate/professional | 1 (7.1%) | 1 (14.3%) | 0 (0.0%) |  |
| **Handedness, n (%)** |  |  |  | 0.266 |
| Right-dominant | 9 (64.3%) | 6 (85.7%) | 3 (42.9%) |  |
| Left-dominant | 5 (35.7%) | 1 (14.3%) | 4 (57.1%) |  |
| **Selected baseline medical characteristics, n (%)** |  |  |  |  |
| Thyroid disease | 4 (28.6%) | 2 (28.6%) | 2 (28.6%) | 1.000 |
| Cardiovascular comorbidity requiring aspirin prophylaxis | 4 (28.6%) | 1 (14.3%) | 3 (42.9%) | 0.559 |
| Gastrointestinal comorbidity | 3 (21.4%) | 2 (28.6%) | 1 (14.3%) | 1.000 |
| Obstructive sleep apnea treated with CPAP | 2 (14.3%) | 1 (14.3%) | 1 (14.3%) | 1.000 |
| Weight-management/metabolic treatment | 2 (14.3%) | 2 (28.6%) | 0 (0.0%) | 0.462 |
| Psychiatric conditions | 6 (42.9%) | 3 (42.9%) | 3 (42.9%) | 1.000 |
| Psychotropic medication use | 5 (35.7%) | 2 (28.6%) | 3 (42.9%) | 1.000 |
| Psychotherapy/behavioral support | 2 (14.3%) | 2 (28.6%) | 0 (0.0%) | 0.462 |

**Abbreviations:** ADHD, attention-deficit/hyperactivity disorder; KBIT-2, Kaufman Brief Intelligence Test–Second Edition; OCD, obsessive-compulsive disorder; SD, standard deviation; tPBM, transcranial photobiomodulation.
**Note:** Psychiatric/behavioral indications included depression, anxiety, ADHD, OCD, or behavioral modification. Psychotropic medication use included sertraline, fluoxetine, guanfacine, and dexamethylphenidate. Psychotherapy/behavioral support was recorded separately when documented as an ongoing non-pharmacologic intervention.

Between-group p values are descriptive only. Continuous variables were compared using Welch’s t-test. Categorical variables were compared using Fisher’s exact test because of the small sample size and sparse cell counts. Ethnicity was not tested because all participants were non-Hispanic/Latino. No multiplicity adjustment was applied.

### Aim 1. Resting-state EEG gamma power

Global resting-state EEG gamma power (35–45 Hz), expressed as log10-transformed mean power, did not significantly increase after active tPBM relative to sham at STF or LTF, despite a descriptive increase in active tPBM and decrease in sham (Tables 4–5; Figure 4).

**Figure 4.**
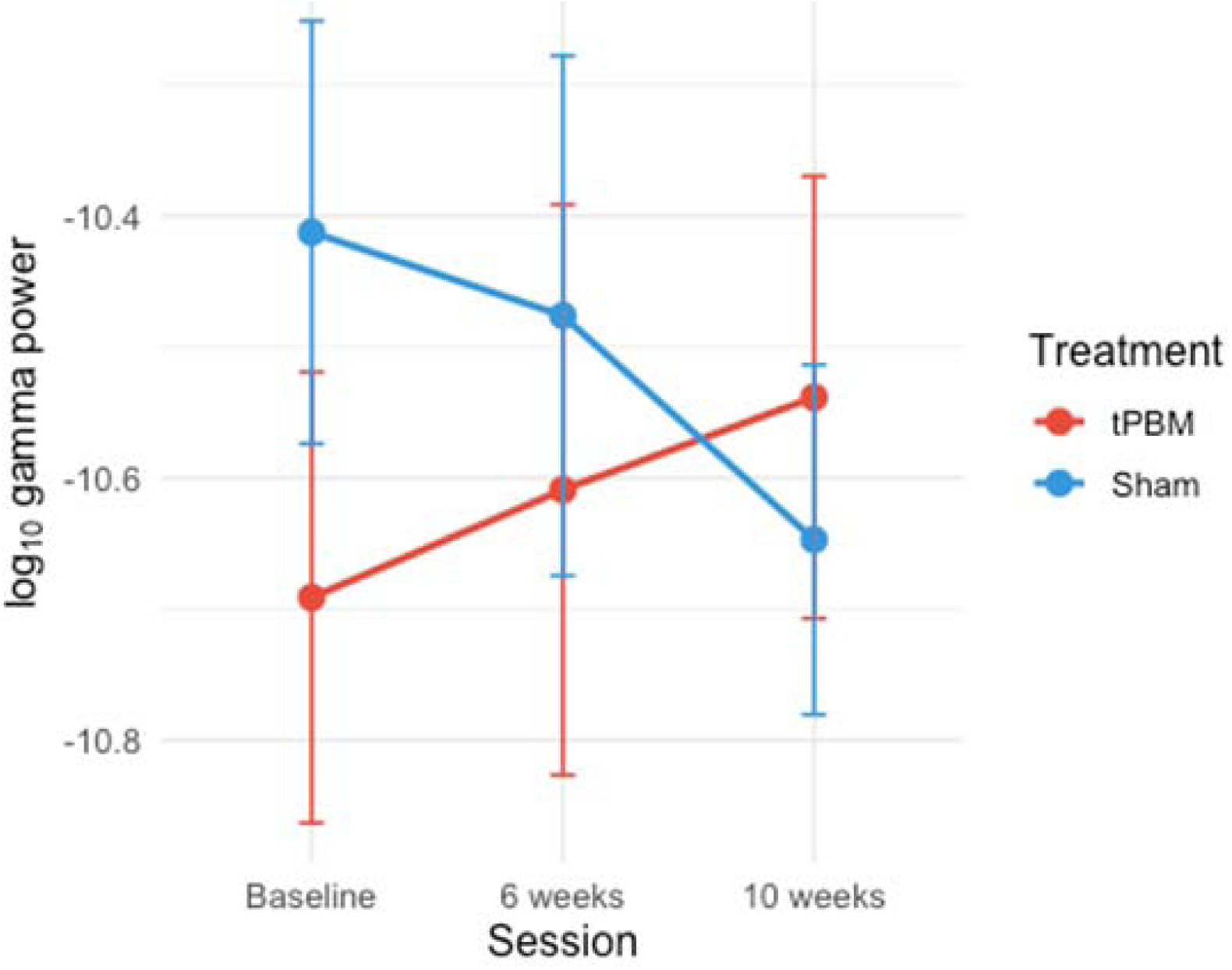
Resting-state EEG gamma power across study visits by treatment group. Mean log10-transformed resting-state EEG gamma power (35–45 Hz) for the active tPBM group and sham group at baseline, post-treatment (6 weeks), and follow-up (10 weeks). Error bars represent standard errors of the mean. The active-treatment group showed a descriptive increase in gamma power over time, whereas the sham group showed a decrease by follow-up; however, between-group treatment effects were not statistically significant.

**Table 4.** Change in resting-state EEG gamma power from baseline.

| Treatment group | Δ STF (post – baseline) | Δ LTF (follow-up – baseline) |
| --- | --- | --- |
| tPBM | 0.0822 ± 0.4499 | 0.1527 ± 0.7704 |
| Sham | –0.0636 ± 0.3229 | –0.2346 ± 0.2940 |

**Table 5.**
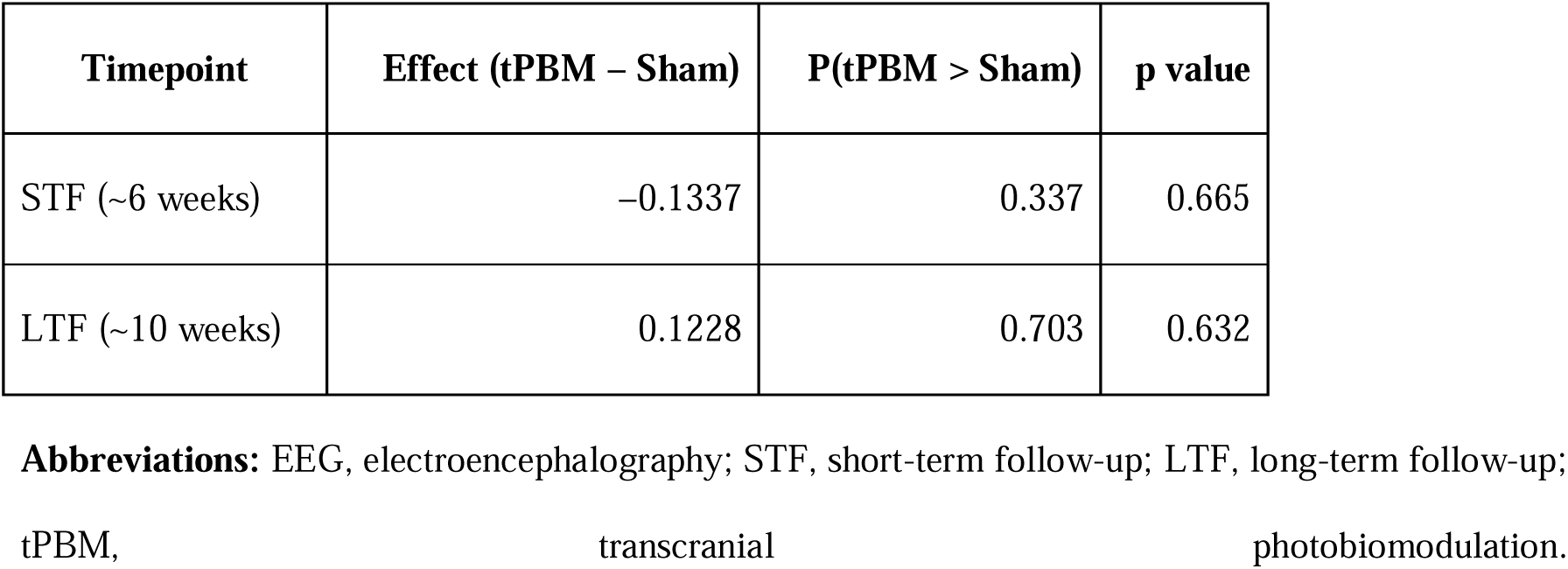
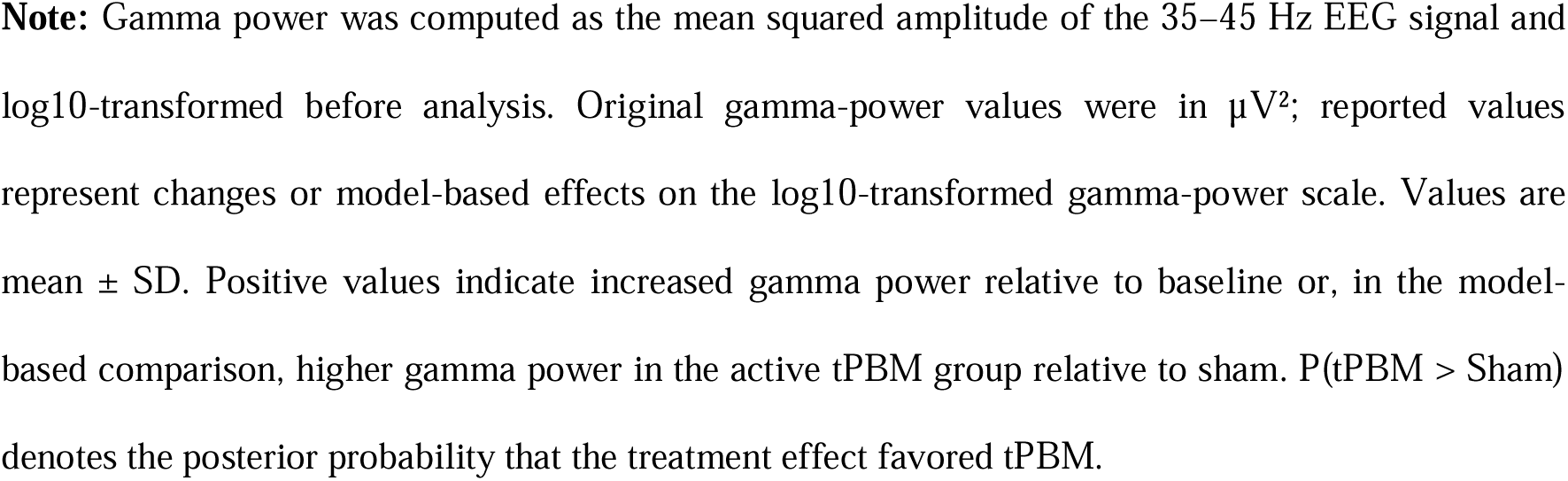
Treatment effects on resting-state EEG gamma power.

| Timepoint | Effect (tPBM – Sham) | P(tPBM > Sham) | p value |
| --- | --- | --- | --- |
| STF (~6 weeks) | –0.1337 | 0.337 | 0.665 |
| LTF (~10 weeks) | 0.1228 | 0.703 | 0.632 |

**Abbreviations:** EEG, electroencephalography; STF, short-term follow-up; LTF, long-term follow-up; tPBM, transcranial photobiomodulation.
**Note:** Gamma power was computed as the mean squared amplitude of the 35–45 Hz EEG signal and log10-transformed before analysis. Original gamma-power values were in $\mu V^2$ ; reported values represent changes or model-based effects on the log10-transformed gamma-power scale. Values are mean ± SD. Positive values indicate increased gamma power relative to baseline or, in the model-based comparison, higher gamma power in the active tPBM group relative to sham. $P(\text{tPBM} > \text{Sham})$ denotes the posterior probability that the treatment effect favored tPBM.

### Aim 2. Language outcomes

#### Wordless Picture Book narrative measures

Grammatical morpheme omissions was the only narrative outcome showing a significant STF between-group difference, favoring active tPBM (median Δ = −1 omission vs +8 omissions in sham; Mann–Whitney p = 0.003). This effect remained significant in the supportive baseline-adjusted model, but was not maintained at LTF. Number of Different Words, intelligibility, and MLU-morphemes were non-significant (Tables 6–7; Figures 5–6).

**Figure 5.**
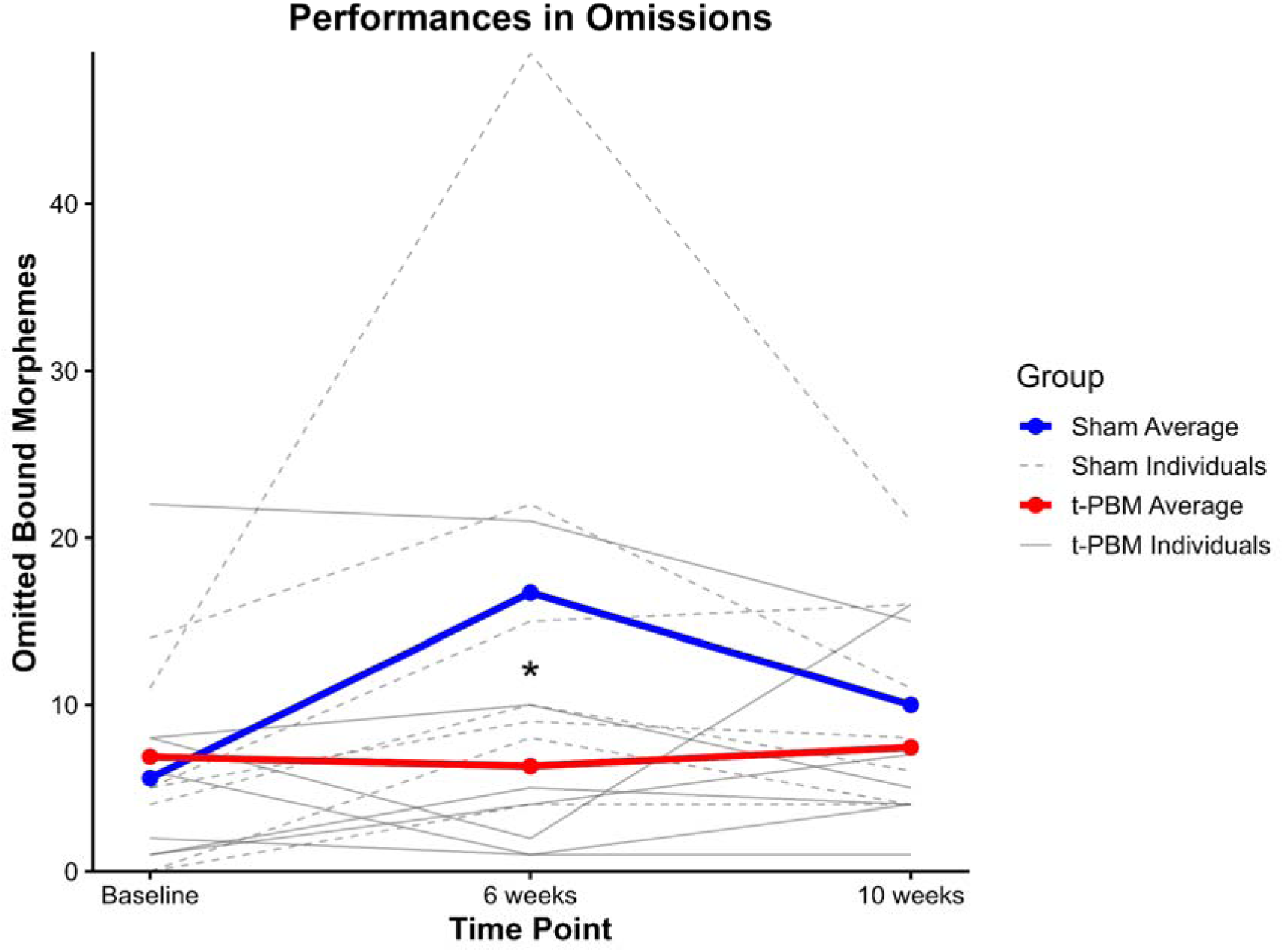
**Grammar Omissions across visits by group.**

**Figure 6.**
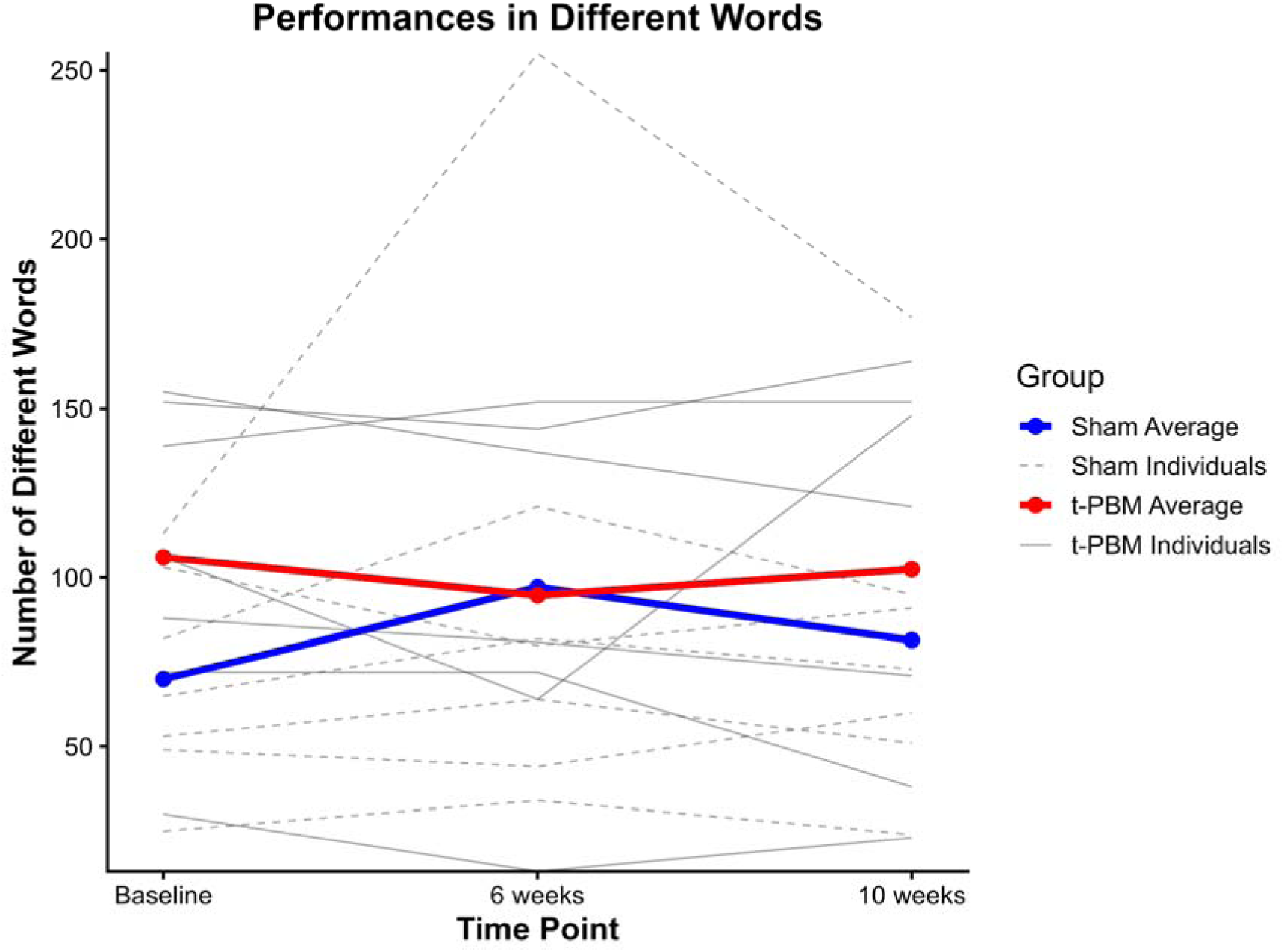
**Number of Different Words across visits by group.**

**Table 6.**
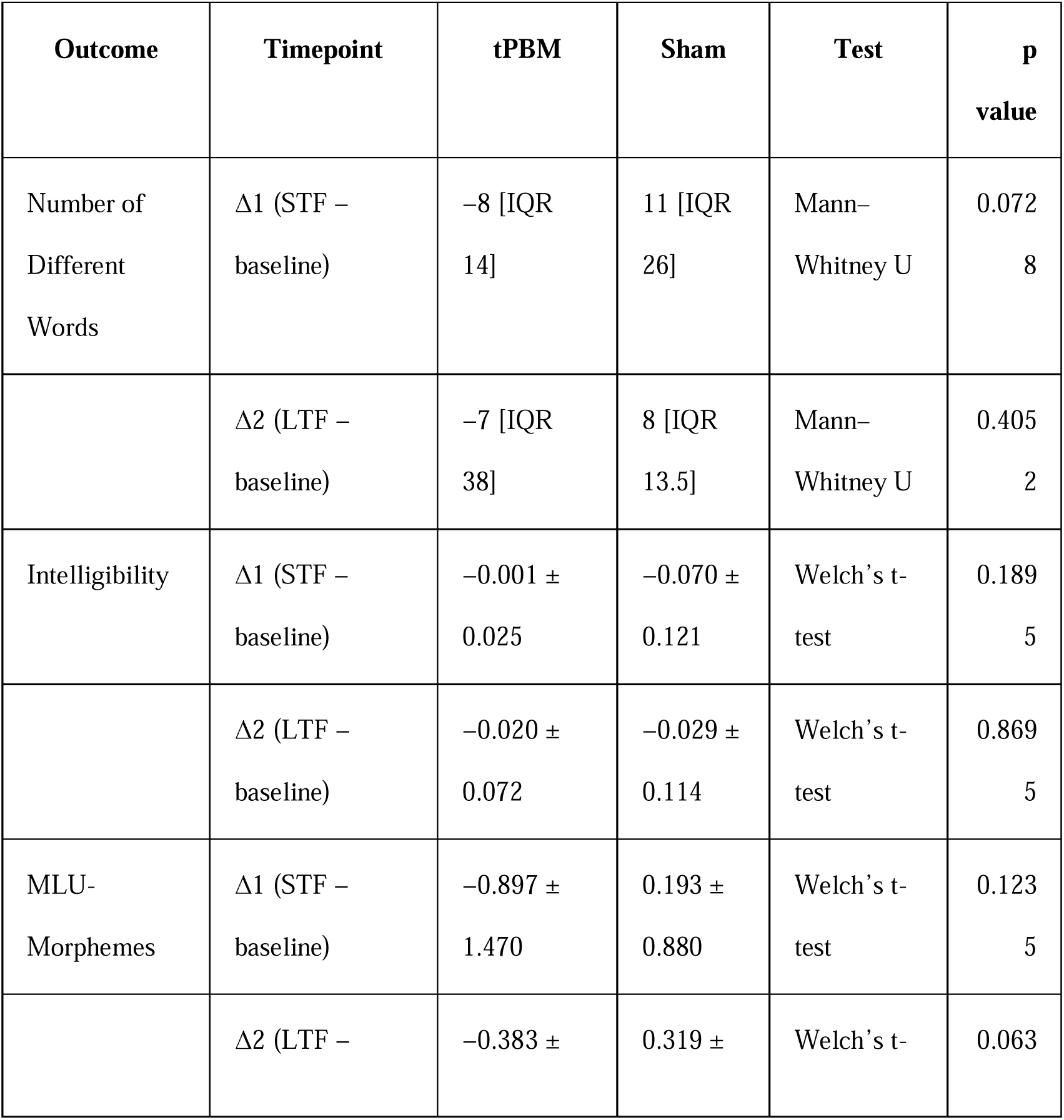
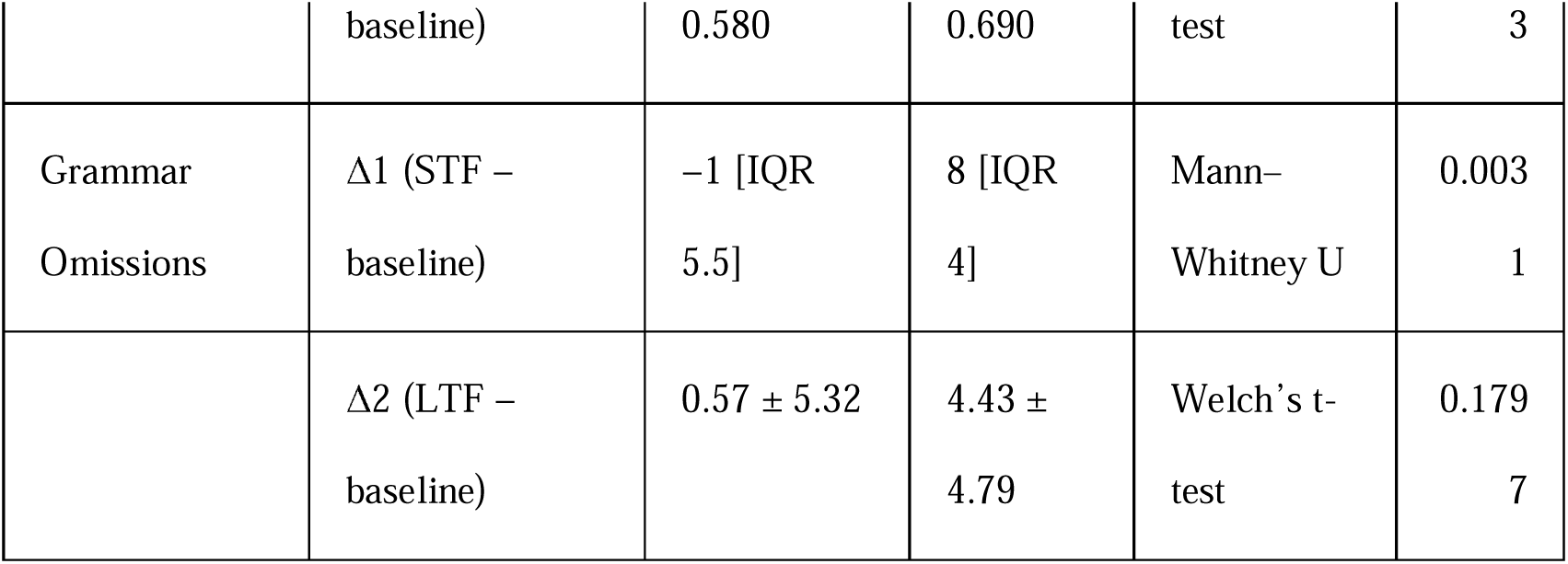
Pre-registered change-score (Δ) analyses in Wordless Picture Book.

**Table 7.**
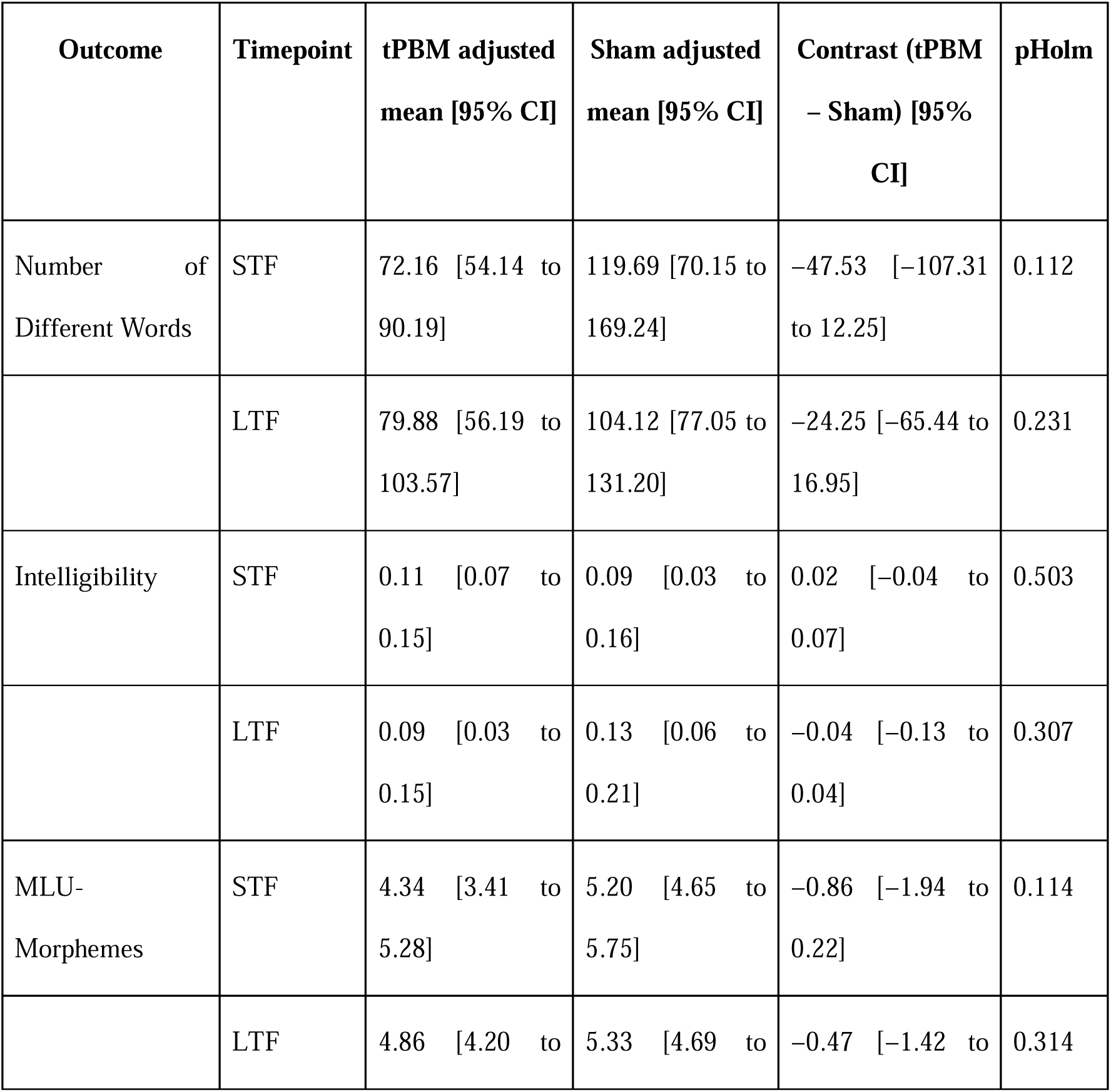
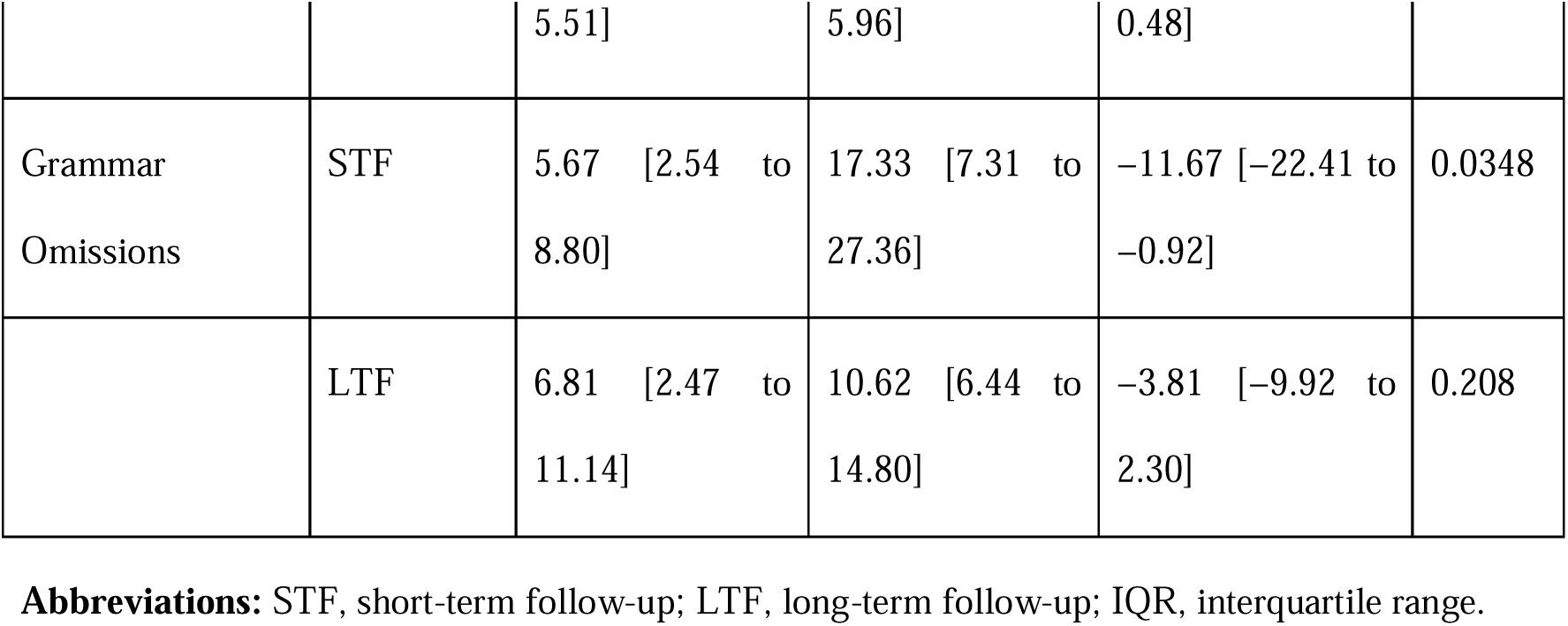
Supportive baseline-adjusted linear mixed-effects models in Wordless Picture Book.

#### RBANS Language Index

Picture Naming was null at STF but favored sham at LTF in both analytic frameworks. Semantic Fluency was non-significant in pre-registered analyses, although the supportive model showed an STF advantage for active tPBM that was not maintained at LTF (Tables 8–9; Figure 7).

**Figure 7.**
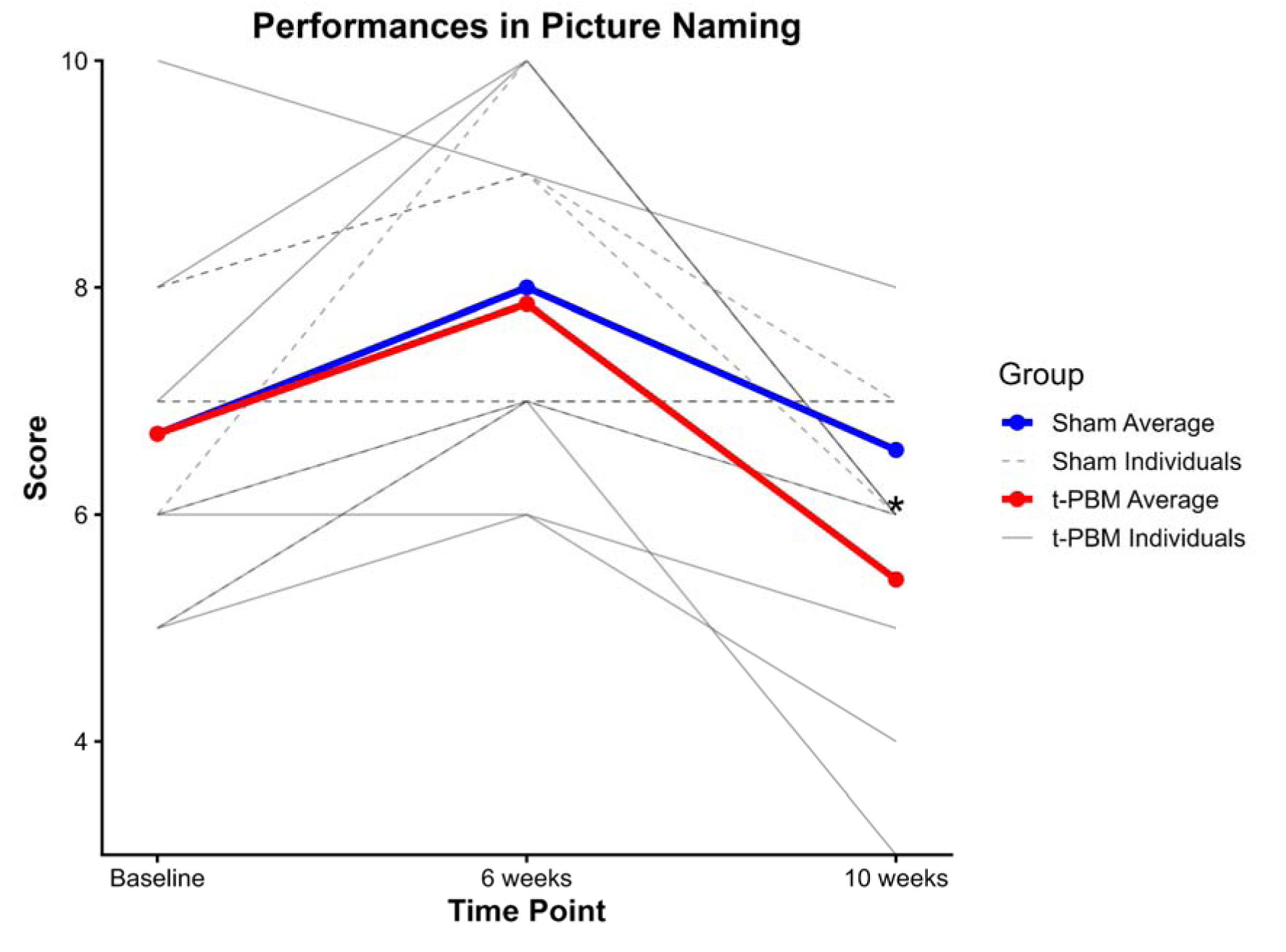
**Picture Naming across visits by group.**

**Table 8.**
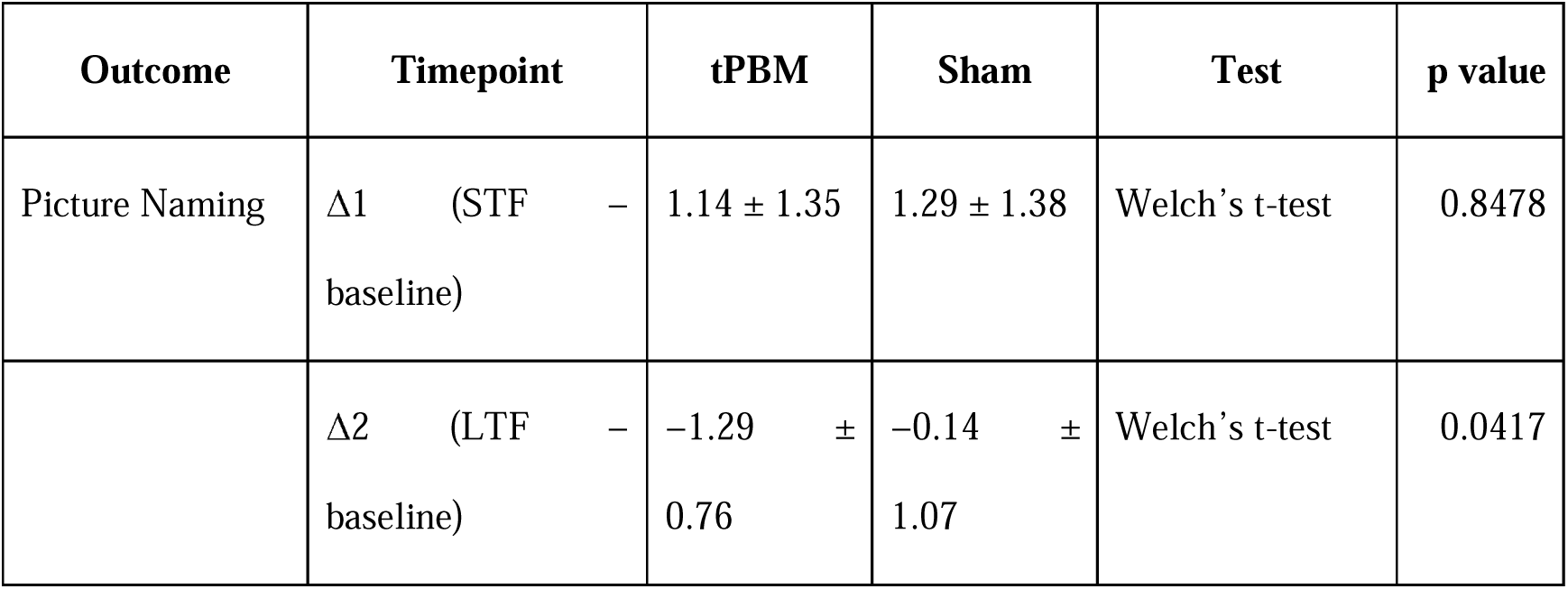
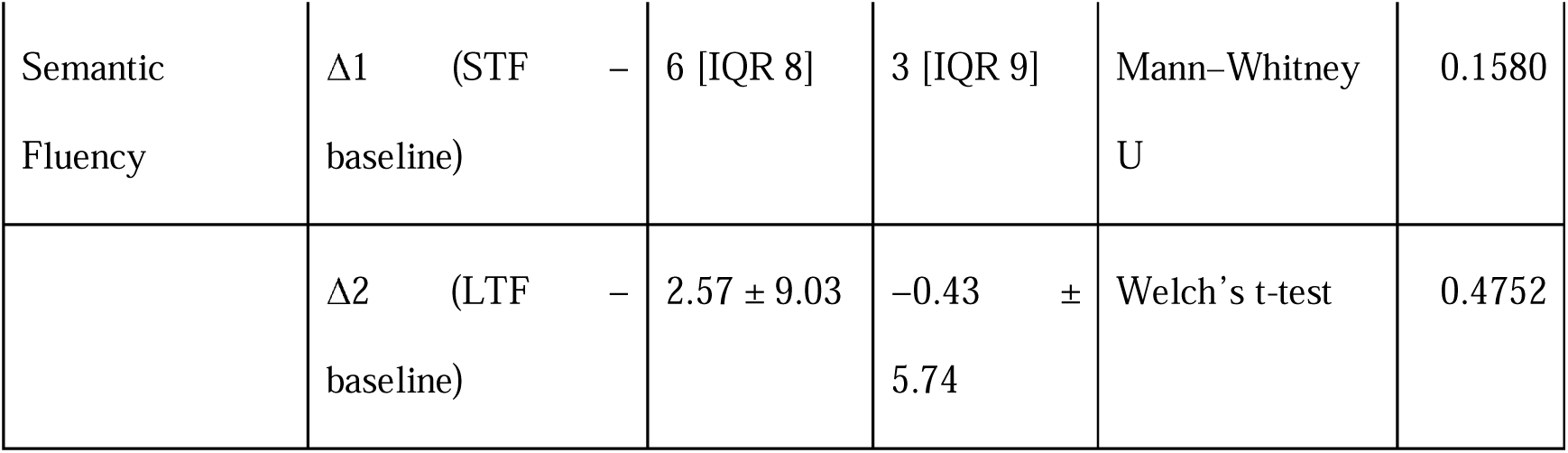
Pre-registered change-score (Δ) analyses in RBANS Language-Index.

**Table 9.** Supportive baseline-adjusted linear mixed-effects models in RBANS Language-Index.

| Outcome | Timepoint | tPBM adjusted mean [95% CI] | Sham adjusted mean [95% CI] | Contrast (tPBM – Sham) [95% CI] | pHolm |
| --- | --- | --- | --- | --- | --- |
| Picture Naming | STF | 7.86 [6.87 to 8.84] | 8.00 [7.03 to 8.97] | –0.14 [–1.53 to 1.24] | 0.833 |
|  | LTF | 5.43 [4.83 to 6.03] | 6.57 [5.99 to 7.15] | –1.14 [–1.98 to –0.31] | 0.00936 |
| Semantic Fluency | STF | 13.62 [11.41 to 15.83] | 9.52 [7.04 to 12.00] | 4.10 [0.80 to 7.39] | 0.0176 |
|  | LTF | 9.76 [7.26 to 12.26] | 7.52 [5.61 to 9.43] | 2.24 [–0.88 to 5.37] | 0.149 |

#### Goldman–Fristoe Test of Articulation–Third Edition (GFTA-3)

GFTA-3 raw articulation error scores showed no significant treatment effects (Tables 10–11; Figure 8).

**Figure 8.**
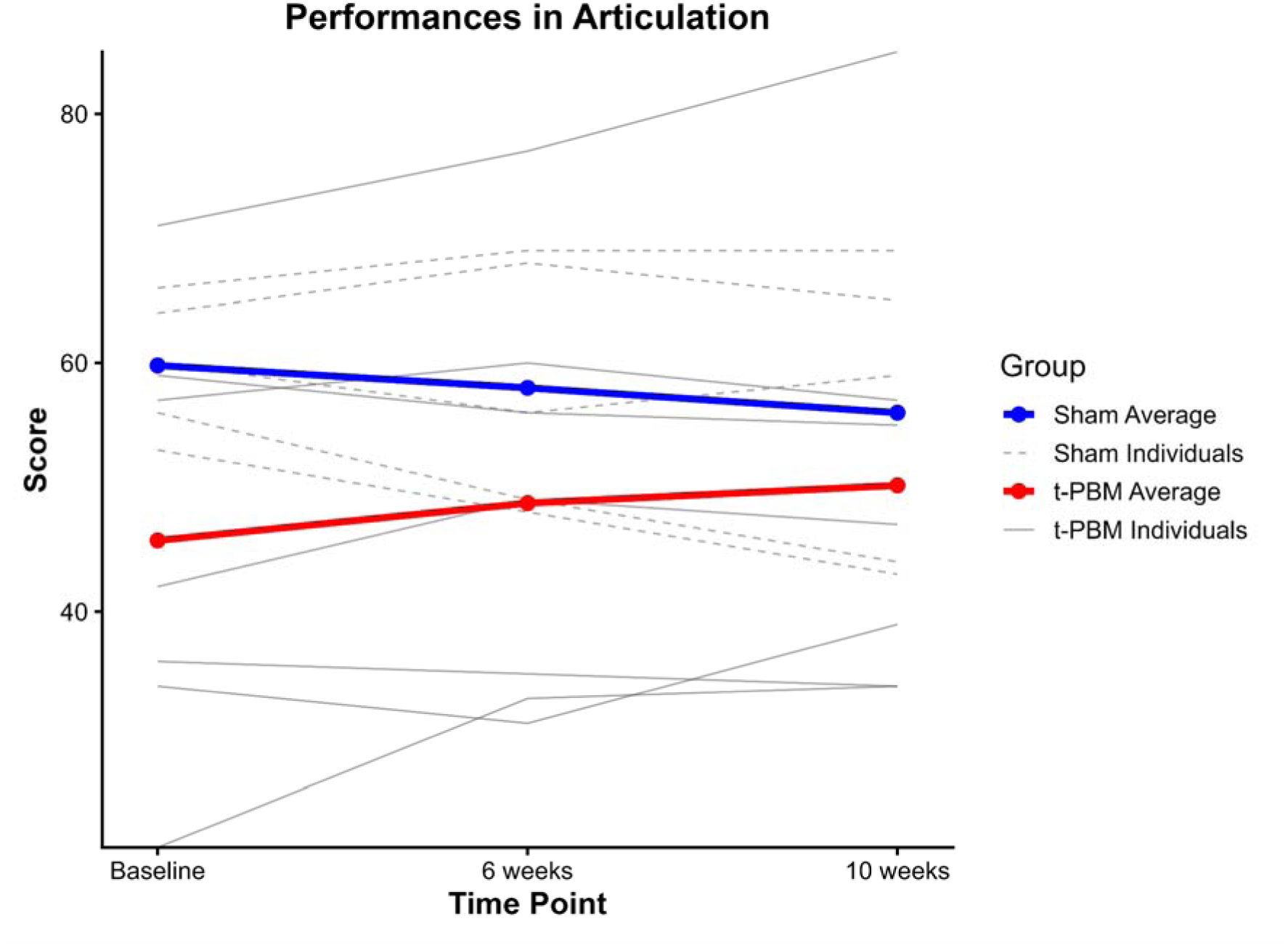
**Language Articulation (GFTA) across visits by group.**

**Table 10.** Pre-registered change-score (Δ) analyses in GFTA.

| Outcome | Timepoint | tPBM | Sham | Test | p value |
| --- | --- | --- | --- | --- | --- |
| GFTA | $\Delta 1$ (STF – baseline) | $3.00 \pm 5.69$ | $-1.80 \pm 4.97$ | Welch's t-test | 0.1533 |
| | $\Delta 2$ (LTF – baseline) | $4.43 \pm 7.04$ | $-3.80 \pm 6.76$ | Welch's t-test | 0.0715 |

**Table 11.** Supportive baseline-adjusted linear mixed-effects models in GFTA.

| Outcome | Timepoint | tPBM adjusted<br>mean [95% CI] | Sham adjusted<br>mean [95% CI] | Contrast (tPBM –<br>Sham) [95% CI] | pHolm |
| --- | --- | --- | --- | --- | --- |
| GFTA | STF | 54.62 [49.36 to<br>59.87] | 49.74 [43.60 to<br>55.88] | 4.88 [–3.56 to 13.31] | 0.228 |
|  | LTF | 56.04 [48.86 to<br>63.23] | 47.74 [40.02 to<br>55.45] | 8.31 [–3.16 to 19.77] | 0.139 |

### Aim 3. Attention and memory outcomes

#### CANTAB

CANTAB findings were inconsistent: RTIFMDMT favored sham at STF only in the pre-registered analysis, PALTEA was null, and MOTML favored active tPBM only in the supportive STF model (Tables 12–13; Figure 9).

**Figure 9.**
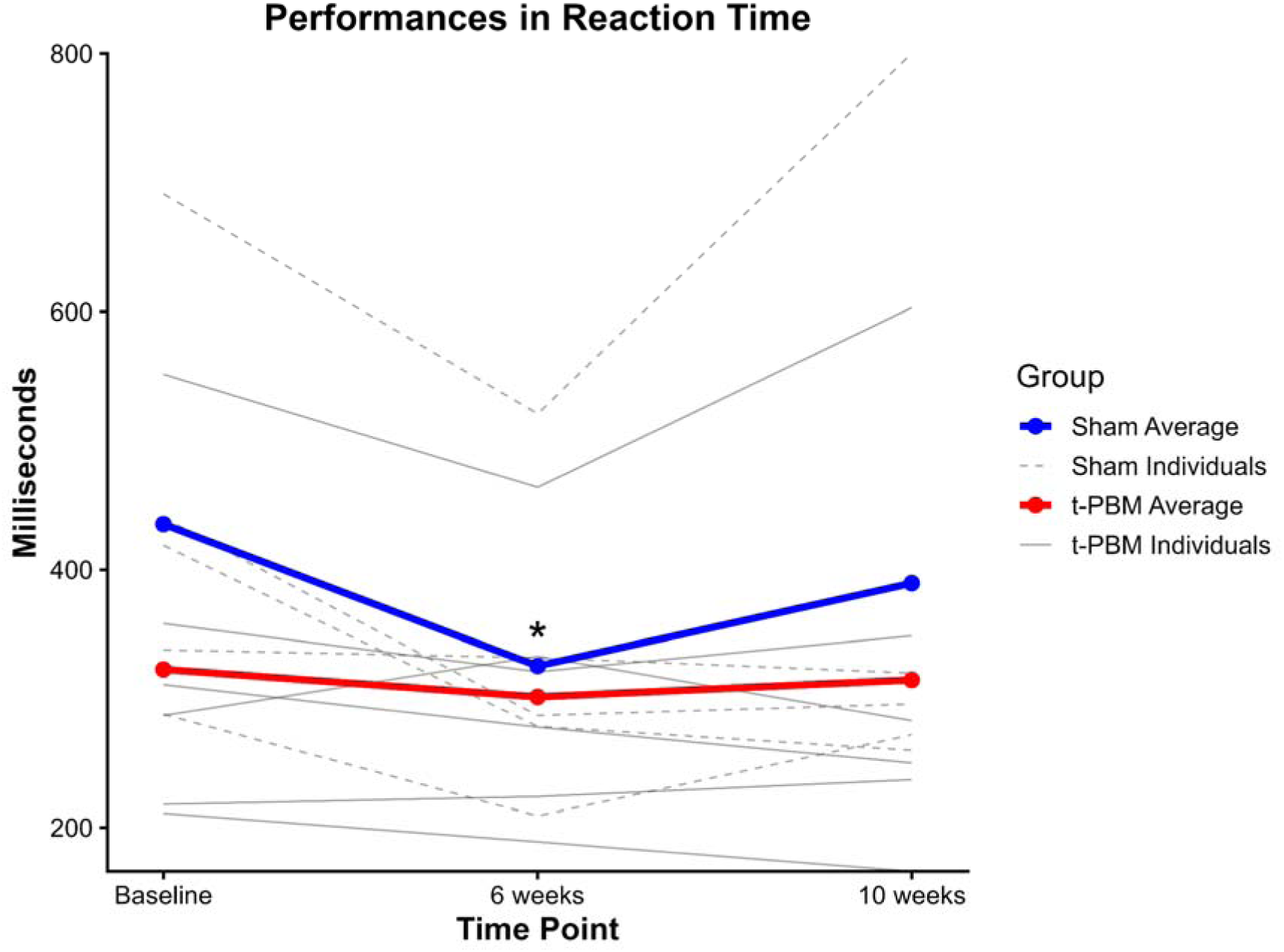
**Reaction Time across visits by group.**

**Table 12.**
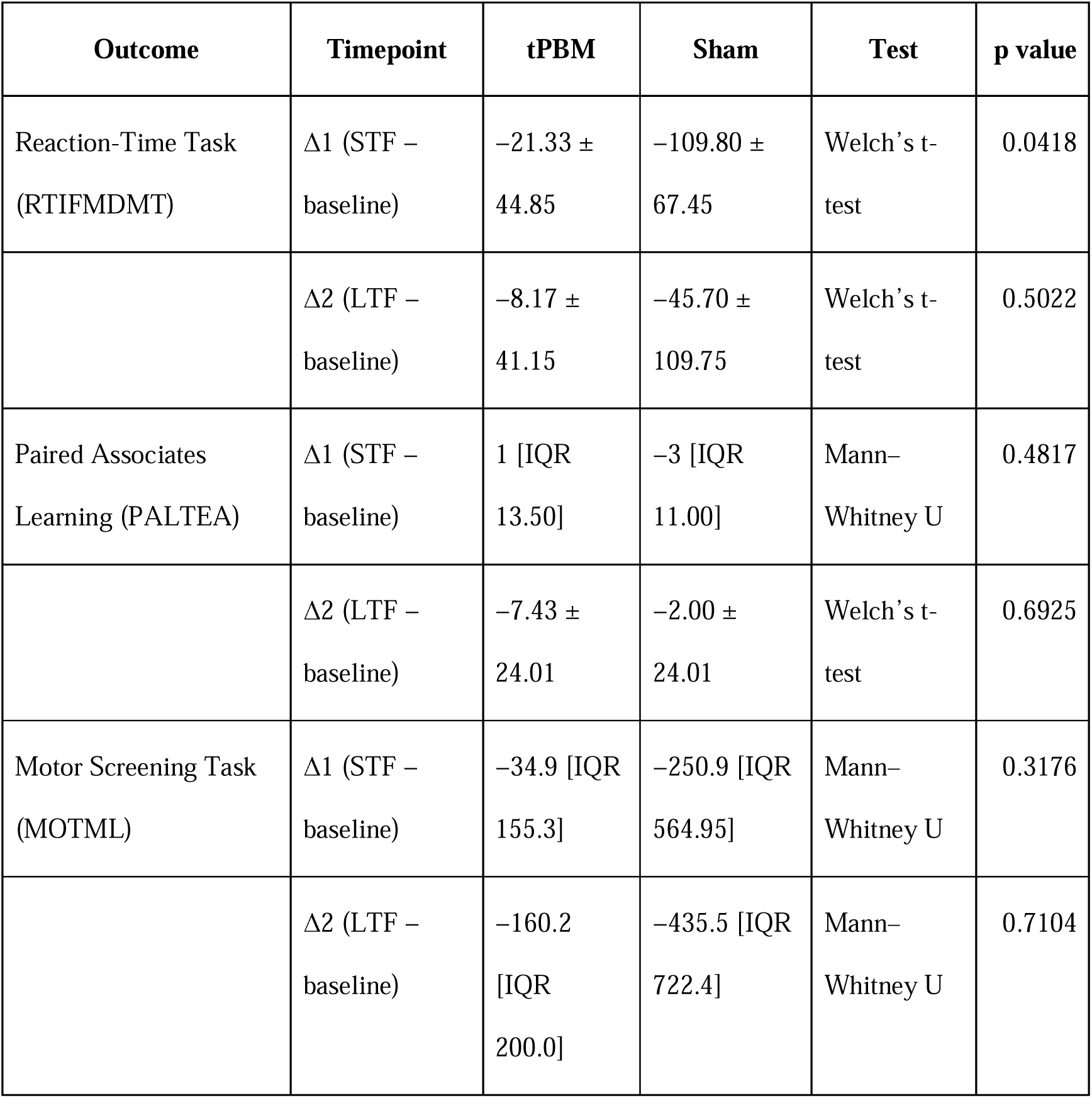
Pre-registered change-score (Δ) analyses in CANTAB.

| Outcome | Timepoint | tPBM | Sham | Test | p value |
| --- | --- | --- | --- | --- | --- |
| Reaction-Time Task<br>(RTIFMDMT) | $\Delta 1$ (STF –<br>baseline) | $-21.33 \pm$<br>44.85 | $-109.80 \pm$<br>67.45 | Welch’s t-test | 0.0418 |
| | $\Delta 2$ (LTF –<br>baseline) | $-8.17 \pm$<br>41.15 | $-45.70 \pm$<br>109.75 | Welch’s t-test | 0.5022 |
| Paired Associates<br>Learning (PALTEA) | $\Delta 1$ (STF –<br>baseline) | 1 [IQR<br>13.50] | -3 [IQR<br>11.00] | Mann–<br>Whitney U | 0.4817 |
| | $\Delta 2$ (LTF –<br>baseline) | $-7.43 \pm$<br>24.01 | $-2.00 \pm$<br>24.01 | Welch’s t-test | 0.6925 |
| Motor Screening Task<br>(MOTML) | $\Delta 1$ (STF –<br>baseline) | $-34.9$ [IQR<br>155.3] | $-250.9$ [IQR<br>564.95] | Mann–<br>Whitney U | 0.3176 |
| | $\Delta 2$ (LTF –<br>baseline) | $-160.2$<br>[IQR<br>200.0] | $-435.5$ [IQR<br>722.4] | Mann–<br>Whitney U | 0.7104 |

**Table 13.**
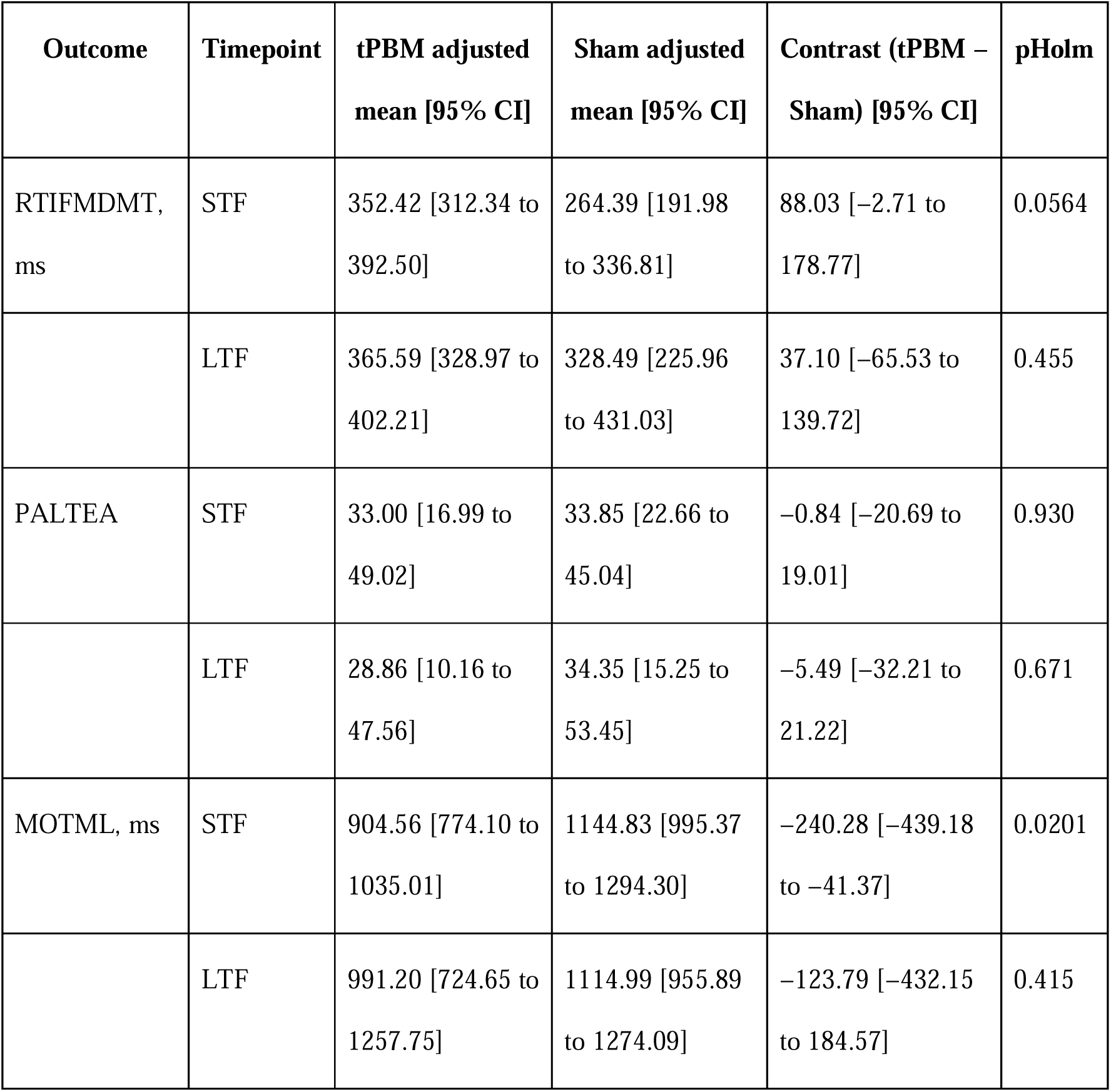
Supportive baseline-adjusted linear mixed-effects models in CANTAB.

#### RBANS Attention and Memory Index

Coding, Digit Span, List Learning, List Recall, and List Recognition showed no significant between-group differences in either analytic framework (Tables 14–15).

**Table 14.**
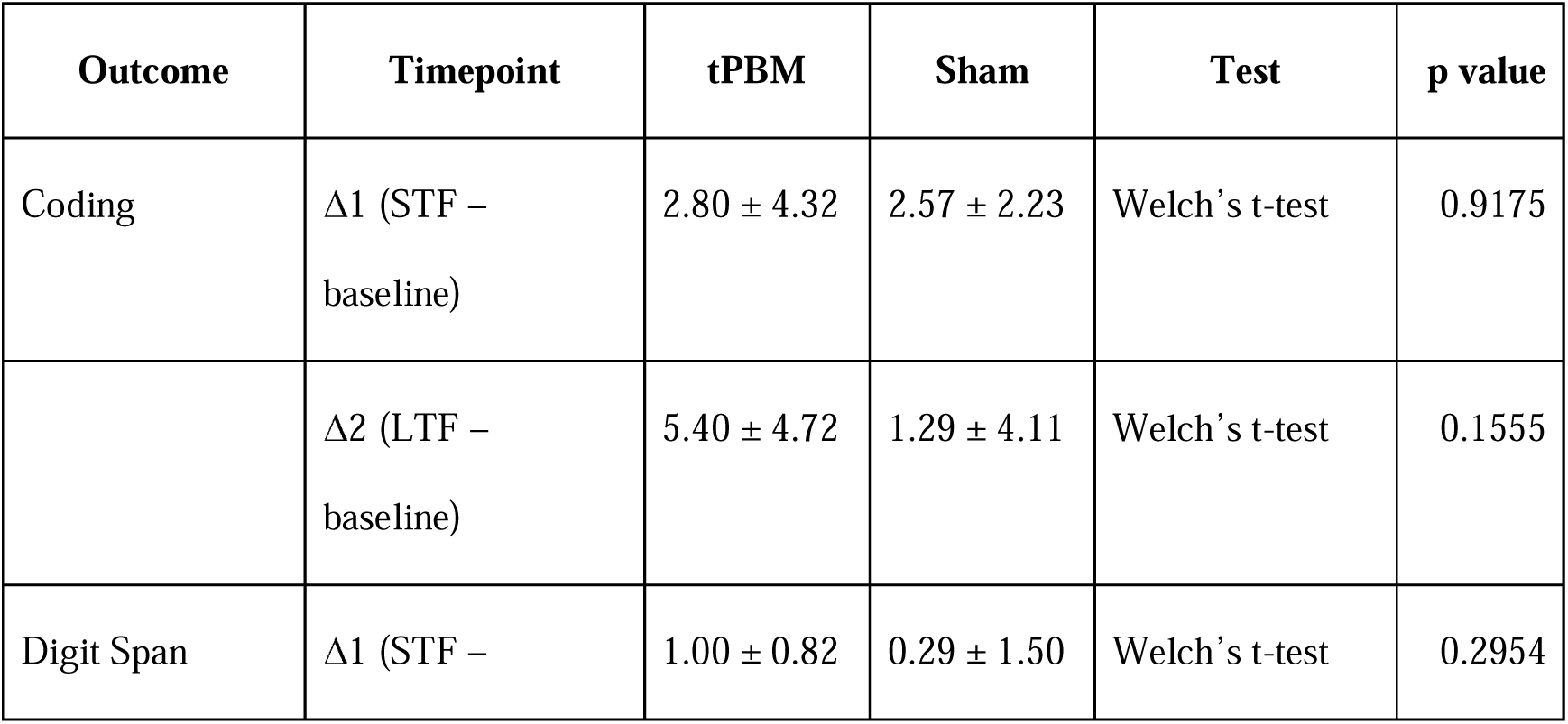
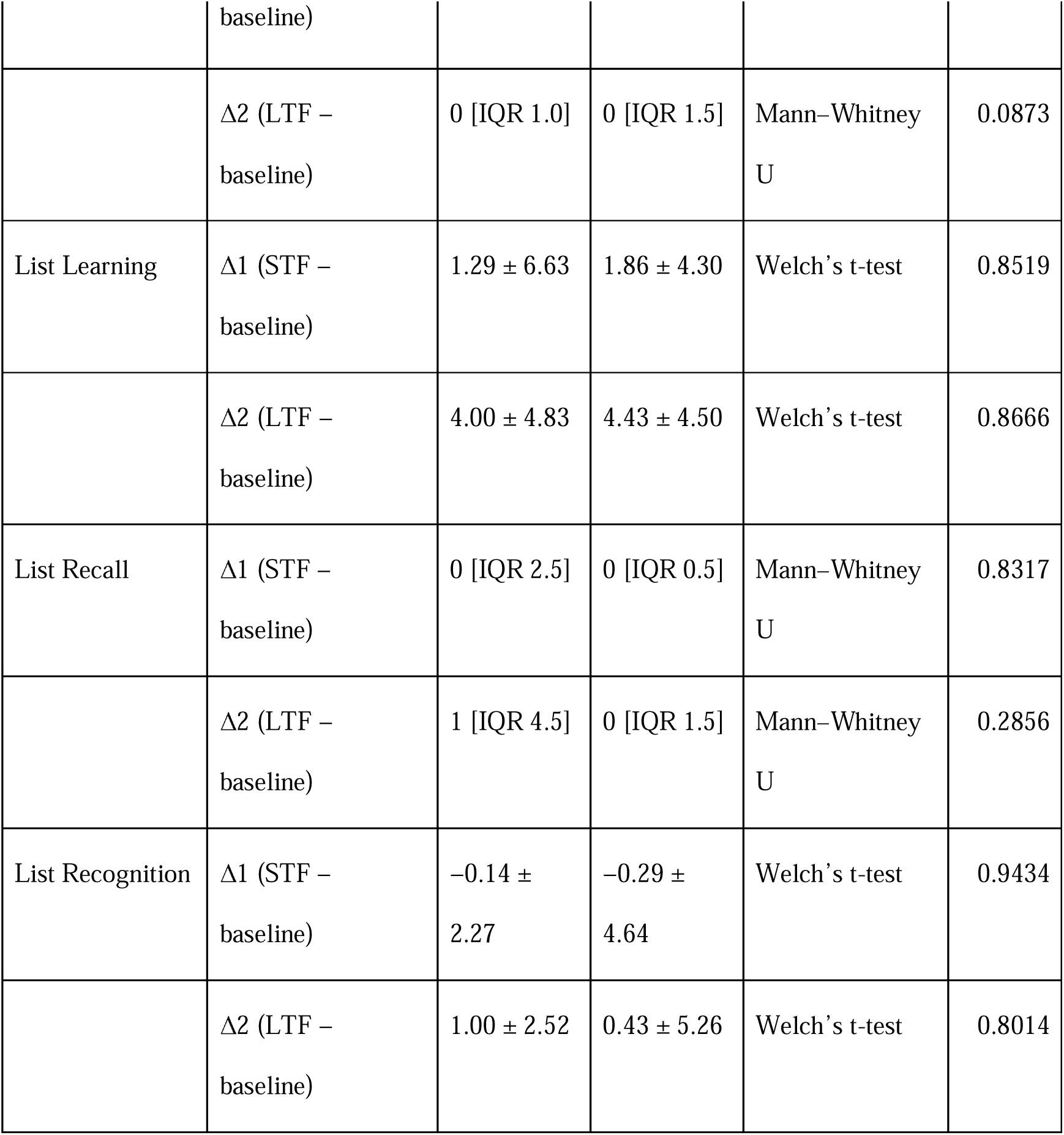
Pre-registered change-score (Δ) analyses in RBANS Memory and Attention-Index.

**Table 15.** Supportive baseline-adjusted linear mixed-effects models in RBANS Memory and Attention-Index.

| Outcome | Timepoint | tPBM adjusted mean [95% CI] | Sham adjusted mean [95% CI] | Contrast (tPBM – Sham) [95% CI] | pHolm |
| --- | --- | --- | --- | --- | --- |
| Coding | STF | 88.83 [–62.13 to 239.78] | 83.74 [–76.94 to 244.43] | 5.09 [–7.51 to 17.68] | 0.393 |
|  | LTF | 90.83 [-59.98 to<br>241.64] | 82.46 [-78.21 to<br>243.12] | 8.37 [-5.51 to<br>22.26] | 0.211 |
| Digit Span | STF | 4.36 [3.69 to<br>5.04] | 3.35 [2.46 to<br>4.24] | 1.01 [-0.14 to<br>2.17] | 0.0817 |
|  | LTF | 3.08 [1.94 to<br>4.21] | 3.92 [3.07 to<br>4.77] | -0.84 [-2.25 to<br>0.56] | 0.224 |
| List Learning | STF | 13.90 [9.34 to<br>18.45] | 13.82 [10.14 to<br>17.50] | 0.08 [-5.67 to<br>5.82] | 0.978 |
|  | LTF | 16.61 [13.32 to<br>19.90] | 16.39 [13.32 to<br>19.46] | 0.22 [-4.19 to<br>4.62] | 0.918 |
| List Recall | STF | 2.99 [0.74 to<br>5.24] | 1.87 [0.92 to<br>2.82] | 1.12 [-1.31 to<br>3.54] | 0.341 |
|  | LTF | 3.70 [1.79 to<br>5.62] | 2.15 [1.12 to<br>3.19] | 1.55 [-0.62 to<br>3.72] | 0.149 |
| List<br>Recognition | STF | 15.23 [12.67 to<br>17.80] | 14.62 [11.10 to<br>18.15] | 0.61 [-3.80 to<br>5.02] | 0.770 |
|  | LTF | 16.38 [14.13 to<br>18.63] | 15.34 [11.38 to<br>19.30] | 1.04 [-3.59 to<br>5.67] | 0.636 |

### Aim 4. Gamma–clinical outcome associations

Exploratory analyses restricted to the active-treatment group examined whether change in resting-state log10 gamma power was associated with change in language and cognitive outcomes. From baseline to post-treatment, correlations were weak for RBANS Language Index (*r* = −0.198), moderate and positive for RBANS List Recall (*r* = 0.410), and strongly negative for RBANS Attention Index (*r* = −0.830; *p* = 0.082). From baseline to follow-up, correlations were modest for RBANS Language Index (*r* = 0.326), RBANS Attention Index (*r* = −0.353), and RBANS List Recall (*r* = −0.428). Given the small active-treatment subgroup, the absence of multiplicity correction, and the lack of statistically significant findings, these analyses do not provide robust evidence that treatment-related gamma changes consistently tracked language or cognitive improvement (Table 16).

**Table 16.** Exploratory correlations between change in EEG gamma power and change in language/cognitive outcomes in the active tPBM group.

| Outcome | Post-treatment r | Follow-up r |
| --- | --- | --- |
| RBANS Language Index | -0.198 | 0.326 |
| RBANS Attention Index | -0.830 | -0.353 |
| RBANS List Recall | 0.410 | -0.428 |

### Exploratory resting-state functional connectivity

Exploratory rs-fMRI analyses did not identify significant treatment effects after correction for multiple comparisons across prespecified networks and regions of interest. At the network level, no robust treatment effects were observed in the Default Mode, Salience, or Central Executive networks. At the regional level, modest positive effect sizes were observed in the angular gyrus and mid-temporal regions, whereas the only nominally significant uncorrected finding was a negative effect in the frontal eye fields, indicating lower functional connectivity in the active-treatment group relative to sham (Table 17). Given the very small imaging subsample and the exploratory nature of these analyses, the rs-fMRI findings should be interpreted cautiously and are best considered hypothesis-generating. Full network- and ROI-level results are provided in Supplementary Figure 1. We acquired dMRI for 6 participants with summary statistics provided in Supplementary Table 2.

**Table 17.**
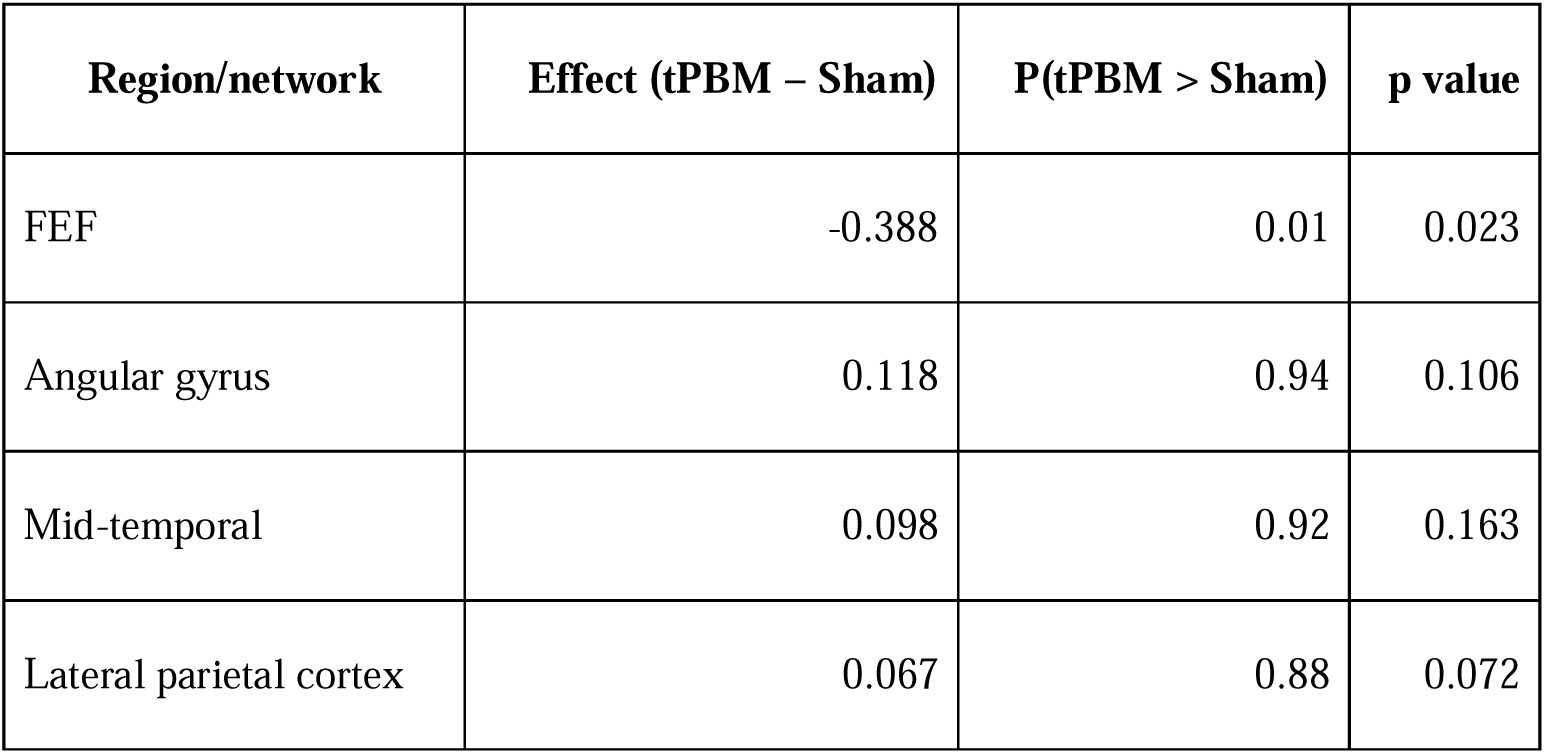
Exploratory resting-state functional connectivity results.

### Safety and tolerability

Both active and sham procedures were well tolerated. Adverse events were reported in all participants, but no serious adverse events occurred and no participant withdrew because of an adverse event. Most events were mild, transient, and local to the treatment site (Table 18).

**Table 18.** Safety and tolerability summary.

| **Safety variable** | **Overall** | **tPBM** | **Sham** |
| --- | --- | --- | --- |
| Participants with $\geq 1$ AE, n/N (%) | 14/14 (100%) | 7/7 (100%) | 7/7 (100%) |
| Total AE entries, n | 248 | 138 | 110 |
| Serious adverse events, n | 0 | 0 | 0 |
| Withdrawals due to AEs, n | 0 | 0 | 0 |
| Participants with local skin reactions, n/N (%) | 13/14 (92.9%) | 6/7 (85.7%) | 7/7 (100%) |
| Participants with headache/head pain, n/N (%) | 4/14 (28.6%) | 2/7 (28.6%) | 2/7 (28.6%) |
| Participants with fatigue/drowsiness, n/N (%) | 6/14 (42.9%) | 3/7 (42.9%) | 3/7 (42.9%) |
| Participants with dizziness, n/N (%) | 4/14 (28.6%) | 2/7 (28.6%) | 2/7 (28.6%) |
| Participants with anxiety/emotional distress, n/N (%) | 6/14 (42.9%) | 3/7 (42.9%) | 3/7 (42.9%) |
| Participants with gastrointestinal symptoms, n/N (%) | 3/14 (21.4%) | 2/7 (28.6%) | 1/7 (14.3%) |

**Abbreviations:** AE, adverse event; tPBM, transcranial photobiomodulation.

## Discussion

This pilot randomized, double-blind, sham-controlled trial did not demonstrate broad efficacy of 6 weeks of 40-Hz near-infrared transcranial photobiomodulation across the prespecified neurophysiological, language, and cognitive domains in adolescents and young adults with Down syndrome. The overall pattern was instead selective and time-limited. The clearest signal was a short-term improvement in grammatical morpheme accuracy during connected speech, whereas broader language, attention, memory, and global resting-state EEG outcomes were largely null. Importantly, findings beyond grammar omissions were not uniformly consistent across analytic frameworks and should therefore be interpreted cautiously.

The most robust language finding was the reduction in grammatical morpheme omissions at short-term follow-up, which was the only language-related outcome replicated across both the pre-registered change-score analyses and the supportive baseline-adjusted mixed-effects models. Given the well-described DS language phenotype, in which expressive language and morphosyntax are disproportionately affected relative to receptive language and some semantic abilities, this pattern may suggest a transient effect on online sentence formulation or morphosyntactic monitoring rather than a generalized enhancement of language output (Martin et al., 2009). Even so, the result remains preliminary and should be viewed as evidence for a potentially sensitive domain-specific endpoint, not for a broad language-facilitating effect (Martin et al., 2009).

By contrast, the absence of reliable between-group differences in number of different words, intelligibility, mean length of utterance in morphemes, and articulation argues against a generalized language benefit under the present protocol. The short-term semantic-fluency advantage is potentially interesting, but it emerged only in the supportive mixed-effects framework and not in the pre-registered primary analyses. Conversely, picture naming favored sham at long-term follow-up in both analytic frameworks. These findings suggest a non-uniform and possibly circuit-specific pattern, in which generative semantic retrieval may be more modifiable than confrontation naming. This interpretation is compatible with lesion and aphasia literature showing that picture naming depends critically on left temporal and inferior parietal systems, and with prior tPBM work in aphasia indicating that naming outcomes may depend strongly on target selection and stimulation parameters (Piai and Eikelboom, 2023; Naeser et al., 2020).

The remaining cognitive findings were similarly mixed. Most RBANS attention and memory outcomes, as well as PAL (Paired Associates Learning), were null across analytic approaches. Reaction-time findings were inconsistent, with the short-term pre-registered RTI (Reaction Time) comparison favoring sham, whereas MOTML (Motor Screening) favored active tPBM only in the supportive mixed model. This pattern does not support a generalized pro-cognitive effect at the tested dose and montage. At most, it raises the possibility of a modest and unstable effect on lower-level sensorimotor efficiency rather than on memory or higher-order attentional performance. Such heterogeneity is plausible given the broader tPBM literature, in which cognitive effects vary substantially across wavelength, fluence, pulsing, montage, exposure schedule, and population studied (Hamblin, 2016; Salehpour et al., 2019; Zomorrodi et al., 2019).

The null result for global resting-state gamma power is central to our interpretation. The biological rationale for this trial was strong, given preclinical evidence linking DS-related gene dosage to altered cortical excitability and impaired gamma oscillations, as well as prior human work suggesting that pulsed tPBM can modulate oscillatory activity (Ruiz-Mejias et al., 2016; Buzsáki and Wang, 2012; Zomorrodi et al., 2019). However, the present EEG outcome was based on a global scalp-averaged 35–45 Hz measure, which may have been insufficiently sensitive to detect focal or state-dependent effects of a montage targeting restricted midline and left perisylvian regions. This concern is reinforced by the fact that EEG acquisition relied on two different systems, which may have introduced additional measurement variability. Accordingly, the absence of a significant global gamma effect weakens the case for robust target engagement under the current protocol, but it does not definitively exclude more localized physiological effects.

The exploratory correlations between gamma change and language/cognitive change should likewise be interpreted cautiously. We did not observe robust evidence that treatment-related changes in gamma consistently tracked changes in language or cognition. However, the large negative association between gamma increase and the RBANS Attention Index, in the active-treatment subgroup, is contrary to our hypothesis. With only seven participants in the active arm, these analyses were necessarily unstable and underpowered, and they should not be taken as strong evidence against a physiology–behavior relationship.

The exploratory rs-fMRI findings also stopped short of demonstrating a reliable network-level effect. The modest, uncorrected signals in angular gyrus and mid-temporal cortex are anatomically interesting because these regions participate in heteromodal association and language-relevant networks, but they remain strictly hypothesis-generating given the extremely small imaging sample and the well-established sensitivity of resting-state functional connectivity estimates to subject motion (Power et al., 2012).

From a translational perspective, the safety profile was reassuring. All randomized participants completed the trial, no serious adverse events occurred, and no participant withdrew because of an adverse event. Most adverse events were mild, transient, and local to the treatment site, with nonspecific symptoms such as headache, fatigue, dizziness, or anxiety occurring in both arms. These findings support the feasibility and tolerability of repeated 40-Hz NIR tPBM in DS, even though efficacy remains unproven. This overall profile is broadly consistent with prior clinical safety studies of tPBM, which reported no serious adverse events and generally mild, transient side effects across repeated-treatment protocols in neuropsychiatric populations (Cassano et al., 2019; Cassano et al., 2022; Coelho et al., 2025).

Several limitations should be emphasized. First, this was a small pilot study and was underpowered to detect anything other than very large effects; notably, the protocol itself indicated that even a sample of 16 would provide 80% power only for an effect size of approximately 1.5, whereas the final study included 14 randomized completers. As a result, null findings should not be interpreted as evidence of no effect, because modest treatment effects may have gone undetected in this small sample. Second, the outcome battery was broad relative to sample size. Because of the high risk of both false-negative and false-positive findings, results emerging only from supportive models should be treated as provisional. Third, treatment exposure was not fully uniform because dose titration below 40 J/cm² was permitted for tolerability and the fifth diode could be omitted in participants with smaller head size. Fourth, only one tPBM dose was tested, largely derived from preliminary clinical observations in younger patients with DS. This dose was characterized by a relatively less penetrating wavelength (870 nm), low average irradiance (22 mW/cm²), target areas partly covered by hair, and less-than-daily sessions over 6 weeks. The use of a single fixed NIR pulse frequency (40 Hz) was also not individualized to participant-specific or region-specific abnormalities in brain oscillations, precluding personalization of stimulation frequency or entrainment strategy. Fifth, mechanistic sensitivity was limited by the use of a global gamma metric, optional imaging, a very small MRI subgroup, and acquisition of EEG on two different systems. Finally, the cohort was young (eligibility 16–35 years; mean age approximately 25 years) and free of dementia, so the Alzheimer-related component of the mechanistic rationale may have been less biologically manifest than in older DS populations (Lott and Head, 2019).

These considerations help define the next step for the field. Future trials should be larger, more mechanistically anchored, and more selective in their endpoint strategy. Rather than treating “language” as a single construct, studies should pre-specify a smaller set of outcomes matched to the hypothesized circuitry, such as connected-speech morphosyntax, semantic retrieval, and confrontation naming, and should pair these with focal physiological measures from the stimulated regions (Martin et al., 2009; Schworer et al., 2022; Piai and Eikelboom, 2023). Dose-finding and montage-optimization studies are also needed, since the human tPBM literature remains highly parameter-sensitive and clinically heterogeneous (Hamblin, 2016; Tedford et al., 2015; Zomorrodi et al., 2019; Naeser et al., 2020). Longer follow-up will be important to determine whether the delayed picture-naming disadvantage observed here was a chance finding or a reproducible off-target effect.

In conclusion, repeated 40-Hz NIR tPBM was feasible and well tolerated in adolescents and young adults with DS, but this study does not support the present protocol as a broad cognitive- or language-enhancing intervention. It does, however, leave open the possibility of selective short-term effects on connected-speech grammar that warrant testing in larger, adequately powered, physiology-informed trials.

## Data Availability

The data that support the findings of this study may be available on request from the corresponding author. The data are not publicly available due to privacy or ethical restrictions.

## Acknowledgements

Computational resources were provided by the Massachusetts Life Sciences Center (MLSC) compute cluster at the Athinoula A. Martinos Center for Biomedical Imaging, Massachusetts General Hospital.

Dr. Nicolas Oreskovic, Dr. Stephanie Santoro, Kavita Krell, Kelsey Haugen, Ayesha Harisinghani, Mikayla Shaffer, Emmy Schumacher, and Amy Torres helped with recruitment from the Massachusetts General Hospital Down Syndrome Program. Benjamin Majewski, resource specialist in the Massachusetts General Hospital Down Syndrome Program, posed for the photographs and modeled the procedures for the study’s informational sheets.

## Declarations

### Ethics approval and consent to participate

The study was approved by the Institutional Review Board of Massachusetts General Hospital/Mass General Brigham **2020P003611**. Written informed consent was obtained from the legal guardian of each participant, and consent/assent was obtained from participants when appropriate.

### Trial registration

ClinicalTrials.gov, **NCT04668001**. Transcranial Photobiomodulation With Near-Infrared Light for Language in Individuals With Down Syndrome (TransPhoM-DS).

## Supplementary Material

Supplementary Material: Clinical outcome measures, analyzed scores, and direction of improvement

**Supplementary Table 1.**
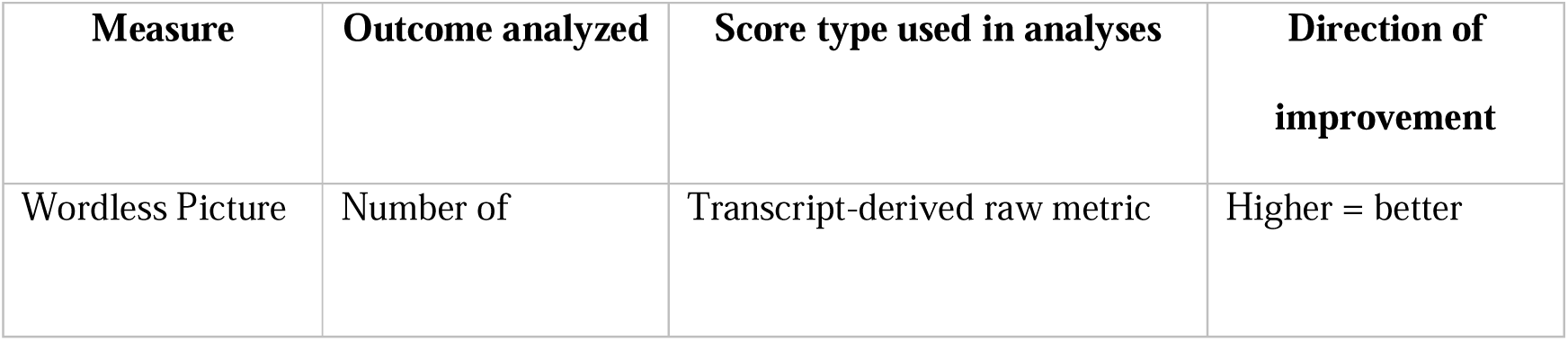
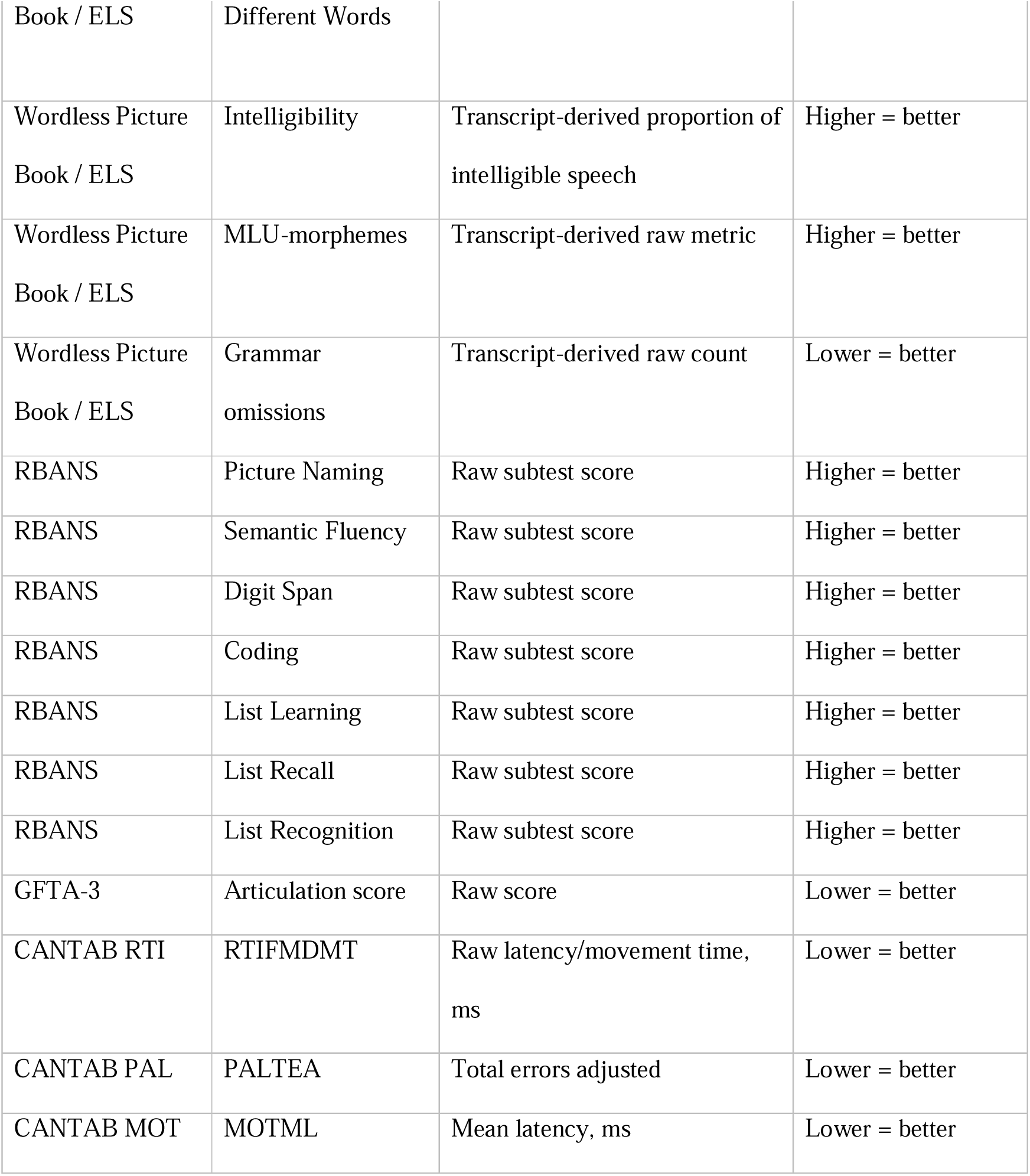
Clinical outcome measures, analyzed scores, and direction of improvement.

Supplementary Material: Exploratory Bayesian analysis of resting-state functional connectivity (rsFC).

**Supplementary Figure 1.**
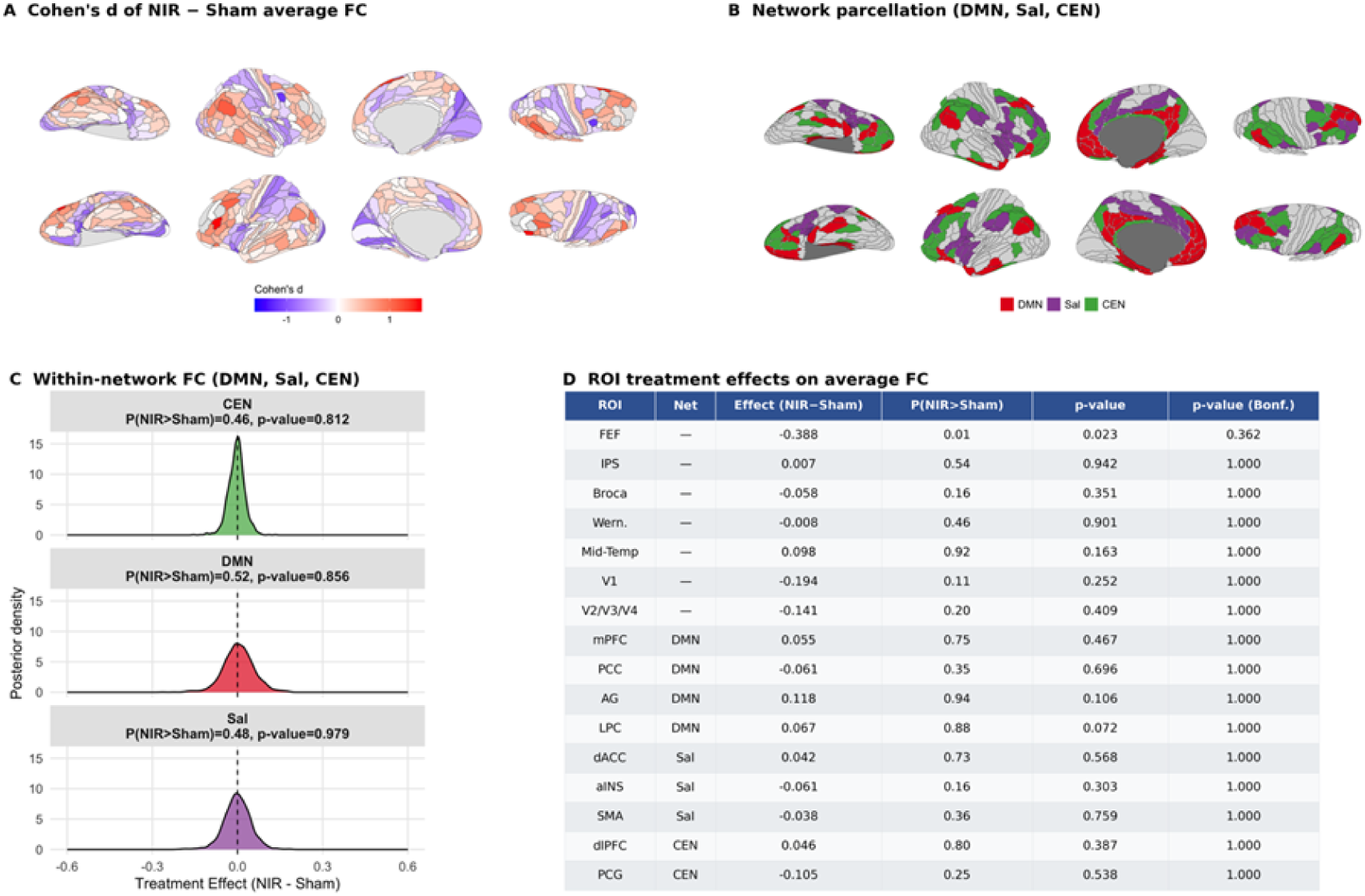
Exploratory Bayesian analysis of resting-state functional connectivity (rsFC). Fisher z-transformed rsFC was compared between the active tPBM/NIR group (n = 3) and sham group (n = 4). (A) Cohen’s d maps for rsFC differences (tPBM − sham). (B) Glasser atlas representation of prespecified Yeo7-derived resting-state networks, including the Default Mode Network (DMN), Salience Network (Sal), and Central Executive Network (CEN). (C) Posterior distributions of treatment effects on within-network rsFC. (D) Region-of-interest treatment effects on average rsFC across prespecified cortical regions, including language-related ROIs. No finding remained significant after correction for multiple comparisons.

Supplementary Material: Diffusion Tensor Imaging Summary

Whole-brain diffusion tensor imaging (DTI) metrics were available for six participants. Because only one participant (113md) belonged to the active tPBM group and all scans exceeded the prespecified motion threshold, no treatment comparisons were performed. For descriptive purposes, whole-brain average values of fractional anisotropy (FA), radial diffusivity (RD), axial diffusivity (AD), and mean diffusivity (MD) are reported in Supplementary Table 1; when multiple sessions were available, values are shown by session.

**Supplementary Table 2.** Whole-brain diffusion tensor imaging (DTI) metrics by participant and session. FA = fractional anisotropy; RD = radial diffusivity; AD = axial diffusivity; MD = mean diffusivity. Values are presented as mean (SD) across white matter voxels. No between-group comparisons were performed because of the very small sample, marked motion burden, and limited representation of the active-treatment group.

| ses | metric | units | 102kd | 107sj | 108je | 112bt | 113md | 118bd |
| --- | --- | --- | --- | --- | --- | --- | --- | --- |
| pre | fa | — | — | — | — | 0.26 (0.21) | — | — |
| | rd | mm<sup>2</sup>/s | — | — | — | $4.9 \times 10^{-4}$ ($4.5 \times 10^{-4}$) | — | — |
| | ad | mm<sup>2</sup>/s | — | — | — | $9.8 \times 10^{-4}$ ($4.5 \times 10^{-4}$) | — | — |
| | md | mm<sup>2</sup>/s | — | — | — | $7.9 \times 10^{-4}$ ($4.3 \times 10^{-4}$) | — | — |
| fu | fa | — | 0.26 (0.22) | 0.23 (0.19) | 0.24 (0.19) | — | — | — |
| | rd | mm<sup>2</sup>/s | $6.7 \times 10^{-4}$ ($4.5 \times 10^{-4}$) | $7.8 \times 10^{-4}$ ($4.6 \times 10^{-4}$) | $7.0 \times 10^{-4}$ ($3.9 \times 10^{-4}$) | — | — | — |
| | ad | mm<sup>2</sup>/s | $9.3 \times 10^{-4}$ ($4.7 \times 10^{-4}$) | $1.1 \times 10^{-3}$ ($4.5 \times 10^{-4}$) | $9.8 \times 10^{-4}$ ($3.9 \times 10^{-4}$) | — | — | — |
| | md | mm<sup>2</sup>/s | $7.6 \times 10^{-4}$ ($4.5 \times 10^{-4}$) | $8.8 \times 10^{-4}$ ($4.4 \times 10^{-4}$) | $7.9 \times 10^{-4}$ ($3.7 \times 10^{-4}$) | — | — | — |
| post | fa | — | 0.26 (0.22) | 0.23 (0.19) | — | — | 0.27 (0.22) | 0.26 (0.21) |
| | rd | mm<sup>2</sup>/s | $6.7 \times 10^{-4}$ ($4.4 \times 10^{-4}$) | $7.7 \times 10^{-4}$ ($4.7 \times 10^{-4}$) | — | — | $7.0 \times 10^{-4}$ ($4.3 \times 10^{-4}$) | $6.7 \times 10^{-4}$ ($3.8 \times 10^{-4}$) |
| | ad | mm<sup>2</sup>/s | $9.4 \times 10^{-4}$ ($4.6 \times 10^{-4}$) | $1.1 \times 10^{-3}$ ($4.6 \times 10^{-4}$) | — | — | $1.0 \times 10^{-3}$ ($4.0 \times 10^{-4}$) | $9.6 \times 10^{-4}$ ($3.5 \times 10^{-4}$) |
| | md | mm<sup>2</sup>/s | $7.6 \times 10^{-4}$ ($4.3 \times 10^{-4}$) | $8.7 \times 10^{-4}$ ($4.5 \times 10^{-4}$) | — | — | $8.0 \times 10^{-4}$ ($4.0 \times 10^{-4}$) | $7.7 \times 10^{-4}$ ($3.5 \times 10^{-4}$) |

